# Risk factors for severe illness and death in COVID-19: a systematic review and meta-analysis

**DOI:** 10.1101/2020.12.03.20243659

**Authors:** Abraham Degarege, Zaeema Naveed, Josiane Kabayundo, David Brett-Major

## Abstract

**Objectives:** This systematic review and meta-analysis synthesized the evidence on the impact of demographics and comorbidities with clinical outcomes of COVID-19, including severe illness, admission to the intensive care unit (ICU), and death.

**Methods:** The PRISMA guidelines were followed to conduct and report this meta-analysis. The protocol is registered in PROSPERO International prospective register of systematic reviews (ID=CRD42020184440). Two authors independently searched literature from PubMed, Embase, Cochrane library and CINHAL on May 6, 2020; removed duplicates; screened titles, abstracts and full text using criteria; and extracted data from eligible articles. A random-effects model was used to estimate the summary odds ratio (OR). Variations among studies were examined using Cochrane Q and I^2^.

**Results:** Out of 4,275 articles obtained from the databases and screened, 71 studies that involved 216,843 patients were abstracted and then, where appropriate, analyzed by meta-analysis. The COVID-19 related outcomes reported were death in 26 studies, severe illness in 41 studies, and admission to ICU in 11 studies. Death was significantly correlated with hypertension (OR 2.60, 95% CI 1.95–3.25, I^2^ = 52.6%, n= 13 studies), cardiovascular disease (5.16, 4.10–6.22, 0.0%, 6), diabetes (2.11, 1.35–2.87, 67.4%, 12), chronic respiratory disease (2.83, 2.14–3.51, 0.0%, 9), cerebrovascular diseases (5.14, 1.08–9.19, 0.0%, 2), male sex (1.34, 1.18 – 1.50, 38.7%, 16), age older than 60 (6.09, 3.53 – 8.66, 95.5%, 6) or 65 years (3.56, 1.21 – 5.90, 18.2%, 6). Severe illness was also significantly associated with hypertension (1.70, 1.30 –2.10, 47.8%, 21), cardiovascular diseases (2.04, 1.01–3.08, 30.6%, 10), diabetes (1.65, 1.23–2.08, 24.9%, 18), male sex (1.35, 1.23 – 1.47, 0.0%, 32) and age at least 60 (4.91, 1.35 – 8.47, 0.0%, 4) or 65 (2.55,1.94 3.17, 24.5%, 9) years. Among hospitalized patients, the odds of admission to ICU was greater in individuals who had cardiovascular diseases (1.36,1.04–1.69, 0.0%, 4), diabetes (1.55, 1.20–1.90, 0.0%, 5) and chronic respiratory disease (1.52, 1.09–1.94, 0.0%, 5) than those who were not having these comorbidities.

**Conclusions:** Older age and chronic diseases increase the risk of developing severe illness, admission to ICU and death among COVID-19 patients. Special strategies are warranted to prevent SARS-CoV-2 infection and manage COVID-19 cases in those with vulnerabilities.

## Introduction

While the numbers of COVID-19 pandemic are a moving target, over 27 million have been infected worldwide, with more than 881 thousand deaths as of September 7, 2020 [1]. Due to potential differences in immunity and genetics, the clinical course of SARS-CoV-2 infection and related death is expected to vary with human health and demography status [2-4]. However, precise factors of importance and their relative roles are not well defined. Study findings on population groups that are more susceptible to severe illness and risk factors for the clinical outcomes in COVID-19 patients remain contradictory [5-9]. While some studies reported an increased risk of developing severe illness and death in COVID-19 patients with hypertension [5-7], cardiovascular diseases [5,7], diabetes [5,6], chronic respiratory diseases [5,6], cancer [6], chronic kidney diseases [6], cerebrovascular disease [6], smoking history [8], ages older than 60 years [7], and male gender [7], a limited number of studies reported lack of correlation between these comorbidities and disease progression or clinical outcomes in COVID-19 patients [9, 10].

Meta-analyses that examined predictors of severe illness and death in COVID-19 patients involved data generated mainly from China and were based on a small number of studies (<15) [11-13]. Although two meta-analyses summarized data based on China, Europe and the USA [14,15], one of these did not examine the effect of some important comorbidities such as cancer, chronic liver disease, and cerebrovascular disease on the risk of admission in ICU or other clinical outcomes [14], the other one searched articles available only in PubMed [15], and both at the time of this submission were not peer-reviewed and are preliminary in nature [14,15]. The relevant literature important to incorporate in systematic review continues to grow, as our collective experience with COVID-19 grows. A better understanding of both striking and subtle variations in patients and their circumstances and their experience with SARS-CoV-2 infection would inform the continued development of improved interventions to reduce morbidity and mortality from COVID-19. Therefore, following PRISMA guidelines, we systematically summarized and assessed literature published before May 6, 2020, and where appropriate to the data, conducted a meta-analysis of risk factors of severe illness, admission to ICU, and death among COVID-19 patients.

## Methods

### Protocol and registration

A protocol developed following the Preferred Reporting Items for Systematic Reviews and Meta-analyses (PRISMA) checklist guided the execution and reporting of this meta-analysis (S1 Table) [16]. The protocol is registered in PROSPERO International prospective register of systematic reviews (ID=CRD42020184440)) [17].

### Inclusion and Exclusion criteria

All retrospective, cross-sectional, and prospective clinical and epidemiological studies except individual case studies (comprehensive inclusion case series allowed), which reported the prevalence or odds of death, severe illness, or admission to ICU and stratified by comorbidities or demographic status among COVID-19 patients were included in the meta-analysis. Unpublished studies and non-peer-reviewed preprints in repositories, case studies with fewer than 10 samples or reports, letters, conference abstracts, protocol, gray literature, review protocols and articles, irrelevant studies (on different topics), and animal or *in vitro* studies were excluded. However, these sources were used to find additional eligible studies.

### Outcome and exposure measures

The primary outcome was death. Secondary outcomes were severe illness and admission to ICU. Severe illness was defined as having one of the following: SpO2 <94%; respiratory rate >30 breaths per minute; lung infiltrates in >50% of lung fields by either plain or computed tomography radiography; arterial partial pressure of oxygen to fraction of inspired oxygen (PaO2/FiO2) <300 mmHg; or, organ dysfunction. Organ dysfunction include respiratory failure as evidenced by mechanical ventilation, myocardial injury as evidenced by the need for catheterization or troponin elevation, renal injury as evidenced by the need for dialysis or a 50% decrease in renal function as assessed by either creatinine rise or decline in glomerular filtration rate, hepatic failure, pulmonary embolus, or stroke/ cerebrovascular accident [18].

The exposure variables were demographic (age, gender, tobacco use) and comorbidities, including hypertension, diabetes, cardiovascular disease, chronic respiratory disease, chronic kidney disease, chronic liver disease, cerebrovascular disease, and cancer.

### Search methods for identification of studies

An article search was conducted in parallel in PubMed, Embase, Cochrane Library, and CINAHL. The search terms were (obesity OR hypertension OR Asthma or nutrition OR age OR gender OR ethnicity OR race OR income OR poverty OR pregnancy OR “Breastfeeding” OR “medical conditions” OR medications OR “chronic diseases” OR influenza or stroke OR HIV OR cancer OR diabetes OR “cardiovascular disease” OR “coronary heart disease” OR “chronic respiratory disease” OR “Sequential Organ Failure Assessment” OR smoking OR “co infection” OR comorbidity OR comorbidities or risk) AND (clinical OR severe OR complications OR mortality OR DEATH) AND (“coronavirus disease 2019” OR “COVID-19” OR “Severe acute respiratory syndrome coronavirus 2” OR SARS-CoV-2 OR “Coronavirus 2” OR “2019 novel coronavirus”). Additional details of the search are available in the supplementary files (S2 Table). After transferring articles searched from the four databases to RefWorks and removing duplicates, the titles and abstracts were screened based on the inclusion and exclusion criteria. Articles approved for full-text review were further screened based on eligibility criteria. The article search and screening processes were conducted by two authors independently. The two authors resolved differences by discussion. A third author was available to mediate disagreement following an independent review.

### Data collection

Data on author, study area/country, study design, sample size, crude or adjusted odds ratio (OR) of death, severe illness or admission to ICU along with 95% confidence interval (CI) among COVID-19 patients with comorbidities *vs* without comorbidities or different demographic status were abstracted from each study. In addition, when OR was not reported, raw data was used to estimate the crude ORs of death, having severe illness or admission to ICU among COVID-19 patients with comorbidities *vs* without comorbidities or different demographic status per study. Two authors abstracted and entered data into the excel sheet independently and compared their results. The final data used for analysis was approved by the two authors upon discussion.

### Quality and risk of bias

The risk of bias and quality of the studies included in this review was evaluated using the Effective Public Health Practice Project tool [19]. The Effective Public Health Practice Project tool uses six criteria: selection bias, design, confounders, blinding, data collection, and withdrawal/dropout, to examine the studies’ quality. Each study’s quality was determined as low, moderate, and high for each of the six criteria using two items for each criterion. The study’s overall quality was determined as high when the study has no weak rating for each of the six characteristics and moderate when the study has one weak rating in one of the six characteristics. The studies were grouped as low quality when the ratings for two or more characteristics were low.

### Data analysis

Percent residual variation among studies due to heterogeneity was estimated using Moran’s I-squared [20]. Statistical significance of the heterogeneity was tested using the Cochran’s Q chi-square test [20]. The odds ratios of the studies combined in the meta-analysis to estimate the summary OR were both adjusted and unadjusted estimates. A random-effect model using the Der Simonian and Laird method was used to estimate the summary ORs. When heterogeneity was high (I^2^>60%), subgroup analysis was performed to estimate the summary ORs after grouping studies by study area/country and study design. Meta-regression was used to explore the sources of heterogeneity at study-level covariates by examining the linear relationship between ORs and study area/country, study design and sample size [21]. A funnel plot that displays the odds ratio estimate against their standard errors was used to evaluate publication bias among the studies included in the meta-analyses [22]. Statistical significance of the asymmetry of the funnel plot was tested using the Egger’s regression test (bias if p<0.1) [23]. A 95% CI and alpha of 0.05 were used to assess the significance of OR.

## Results

The initial search of articles from the databases using the keywords resulted in 4275 articles (PubMed=1986, Embase= 2006, CINAHL =224, and Cochrane Library = 59) out of which 1527 were duplicates (Fig 1). Of the non-duplicate 2748 articles, 2444 were ineligible after screening the titles and abstracts, and 233 articles were excluded after full-text review. This resulted in 71 articles appropriate for inclusion in the systematic review [5-10, 24-89], and 60 of them were also included in the meta-analysis. The majority of the studies were conducted in China (n=53), and the others from South Korea (n=2), Iran (n=1), Europe (n=6) and USA (n=9) (S1 Table 3). Study designs included retrospective (n=42), cross-sectional (n=25), and prospective (n=4). Clinical outcomes reported were death in 34 studies, severe illness in 45 studies, and admission to ICU in 11 studies. Exposure variables examined in the studies included hypertension, cardiovascular disease, diabetes, chronic respiratory disease, cancer, chronic kidney disease, chronic liver disease, cerebrovascular disease, smoking, age and sex.

**Fig 1.**
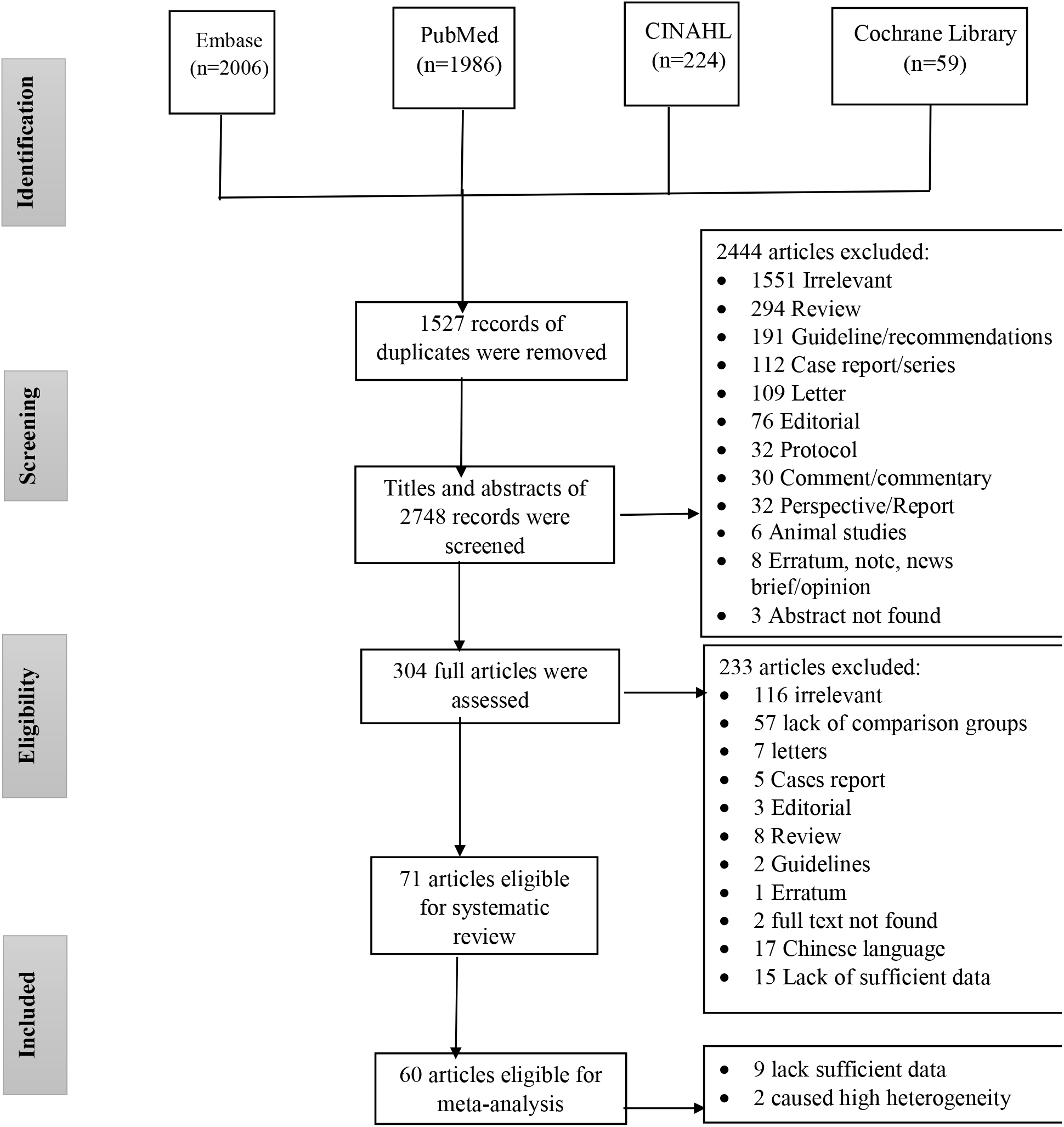
PRISMA flow diagram. Number of articles retrieved from databases, screened, excluded, and included.

### Hypertension

Out of 71 studies approved for inclusion in this review, 38 examined the correlation of hypertension with death, severe illness, and admission to ICU among COVID-19 patients. Out of the 38 studies, 36 included in the meta-analyses. Figure 2 shows summary estimates of the odds ratio of death (*vs* survival: n=13 studies), severe illness (*vs* moderate or mild: n=21 studies), and admission to ICU (*vs* non-ICU: n=4 studies) among hypertensive *vs* non-hypertensive patients. While some studies reported increased odds of death (n=9 studies) and severe illness (n=10 studies) among hypertensive patients, others reported a lack of correlation between hypertension and death (n=4 studies) or severe illness (n=11 studies). A summary analysis of pooled data from these studies showed moderate to high heterogeneity. Though, there were greater odds of death (OR 2.60, 95% CI 1.95–3.25, I^2^ = 52.6%, number of studies (n)= 13, number of participants (N)=53,222) and severe illness (OR 1.70, 95% CI 1.30 –2.10, I^2^ = 47.8%, n=21, N=6,172) among hypertensive as compared to non-hypertensive patients. The odds of admission to ICU were comparable between those who were hypertensive *vs* those who were not hypertensive (OR 1.20, 95% CI 0.30 –2.18, I^2^ = 79.2%, n=5, N=1,832).

**Fig 2.**
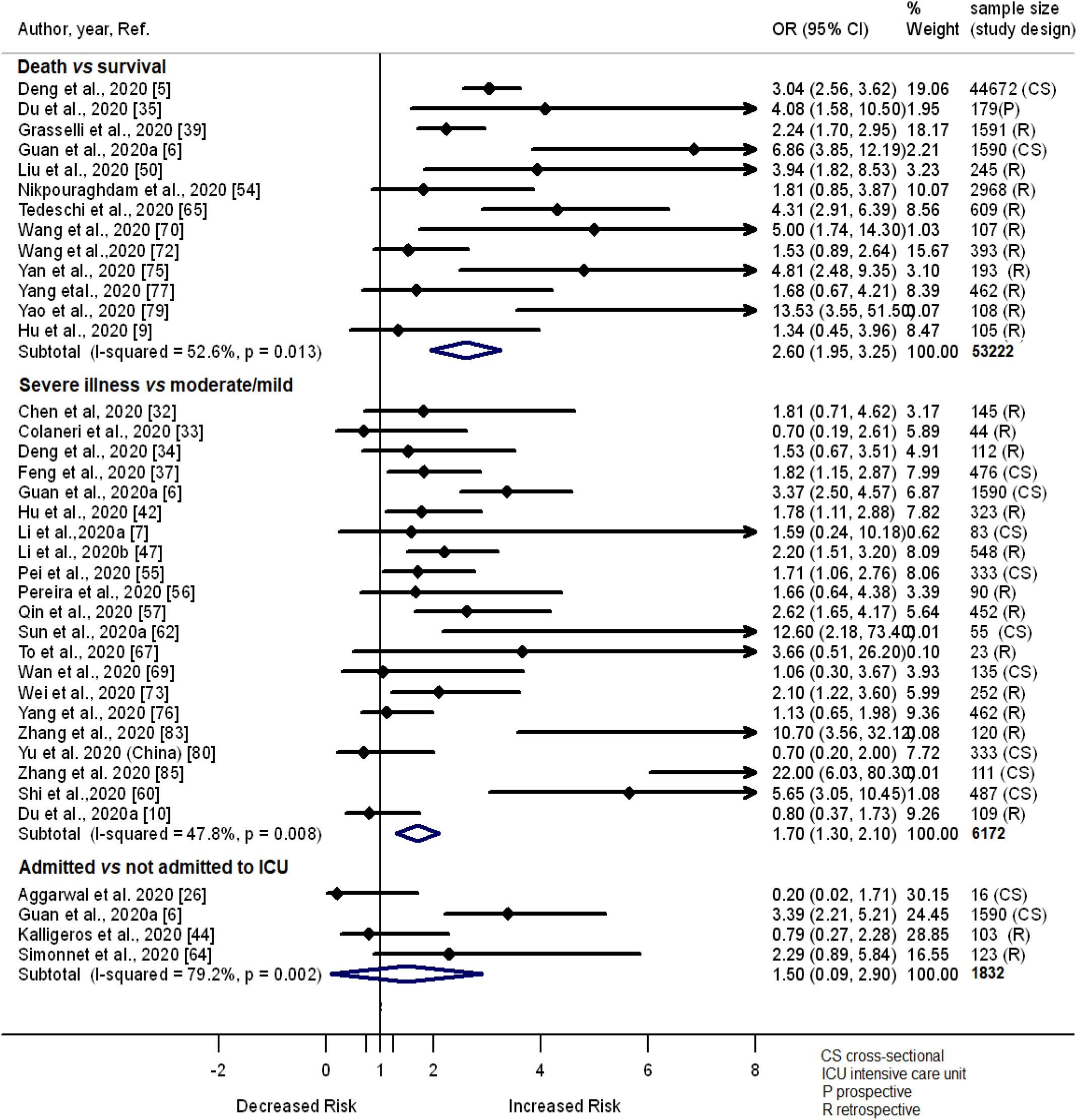
Forest plot showing the relationship of hypertension with the odds of death, severe illness and admission in ICU among COVID-19 patients.

### Cardiovascular disease

Twenty studies examined the nature of the relationship of cardiovascular disease with the odds of developing death (n=7), severe illness (n=11) and admission to ICU (n=5) among COVID-19 patients. Out of the twenty studies, the majority showed higher odds of death (n=6) and severe illness (n=8) among COVID-19 patients with cardiovascular disease (Fig 3). The remaining 1 out of 7 studies showed a lack of association between cardiovascular disease and death, and 3 out of 11 showed a lack of association between cardiovascular disease and the odds of developing severe illness among COVID-19 patients. A meta-analysis of the studies showed higher odds of death (OR 5.16, 95% CI 4.10–6.22, I^2^ = 0.0%, n=6, N=47,134), severe illness (OR 2.04, 95% CI 1.01–3.08, I^2^ = 30.6%, n=11, N=48,535) and admission to ICU (OR 1.36, 95% CI 1.04–1.69, I^2^ = 0.0%, n=5, N=8,346) among COVID-19 patients who had cardiovascular disease as compared to those without this health problem (Fig 3).

**Fig 3.**
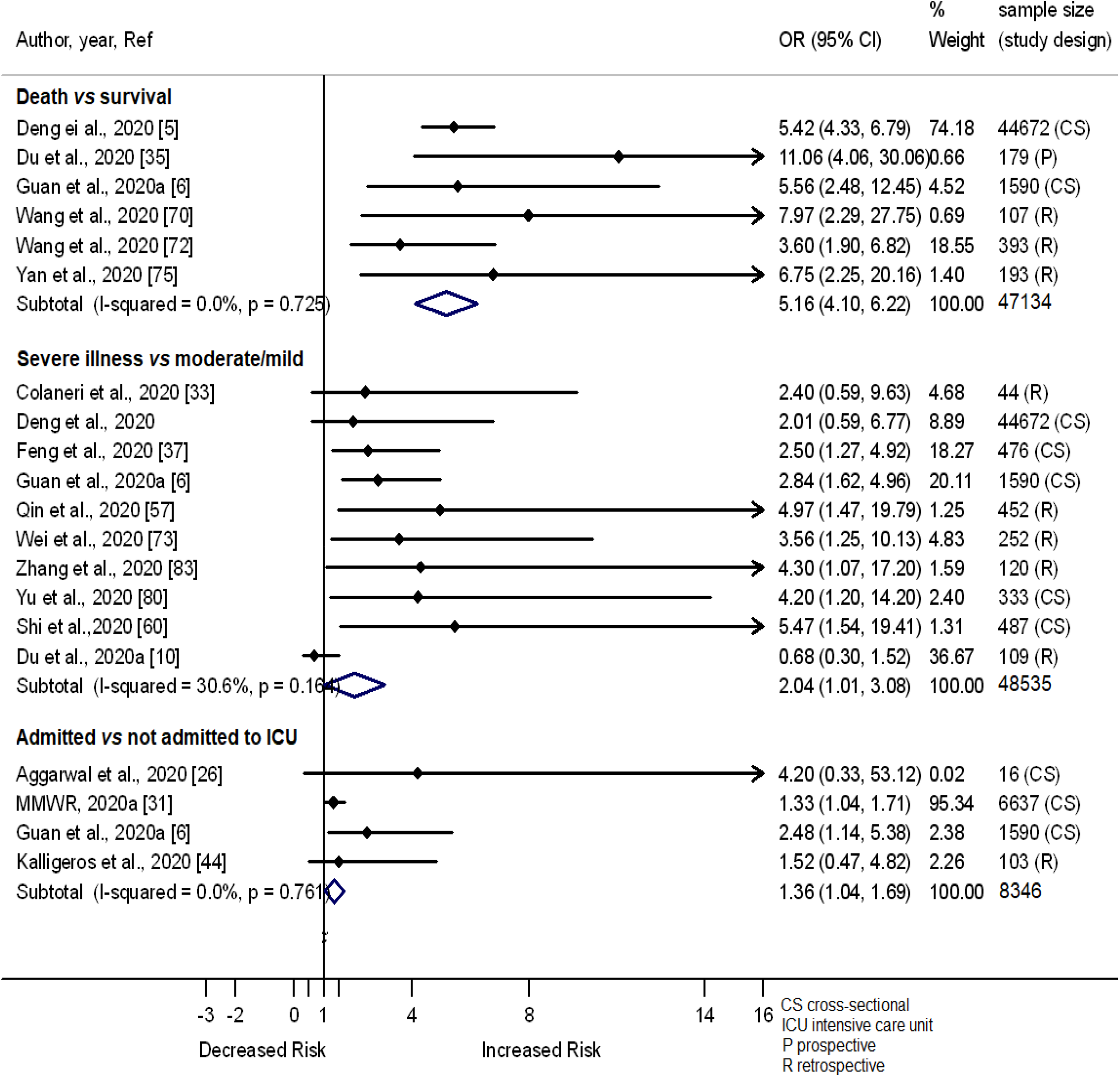
Forest plot showing the relationship of cardiovascular disease with the odds of death, severe illness and admission in ICU among COVID-19 patients.

### Diabetes

Diabetes also is posited to be linked with the risk of developing severe illness and death among COVID-19 patients. Twelve studies examined if having diabetes is correlated with the odds of death among COVID-19 patients, half of which reported increased odds of death in COVID-19 patients with diabetes. Correlation of having diabetes with the odds of developing severe illness and the odds of admission to ICU was assessed in 18 (eight reported increased odds) and five (two reported increased odds) studies, respectively. A summary analysis of these studies showed greater odds of death (OR 2.11, 95% CI 1.35–2.87, I^2^ = 67.4%, n=12, N=59281), severe illness (OR 1.65, 95% CI 1.23–2.08, I^2^ = 24.9%, n=18, N=5811) and admission to ICU (OR 1.55, 95% CI 1.20–1.90, I^2^ = 0.0%, n=5, N=8469) among patients with diabetes than those who had no diabetes (Fig 4).

**Fig 4.**
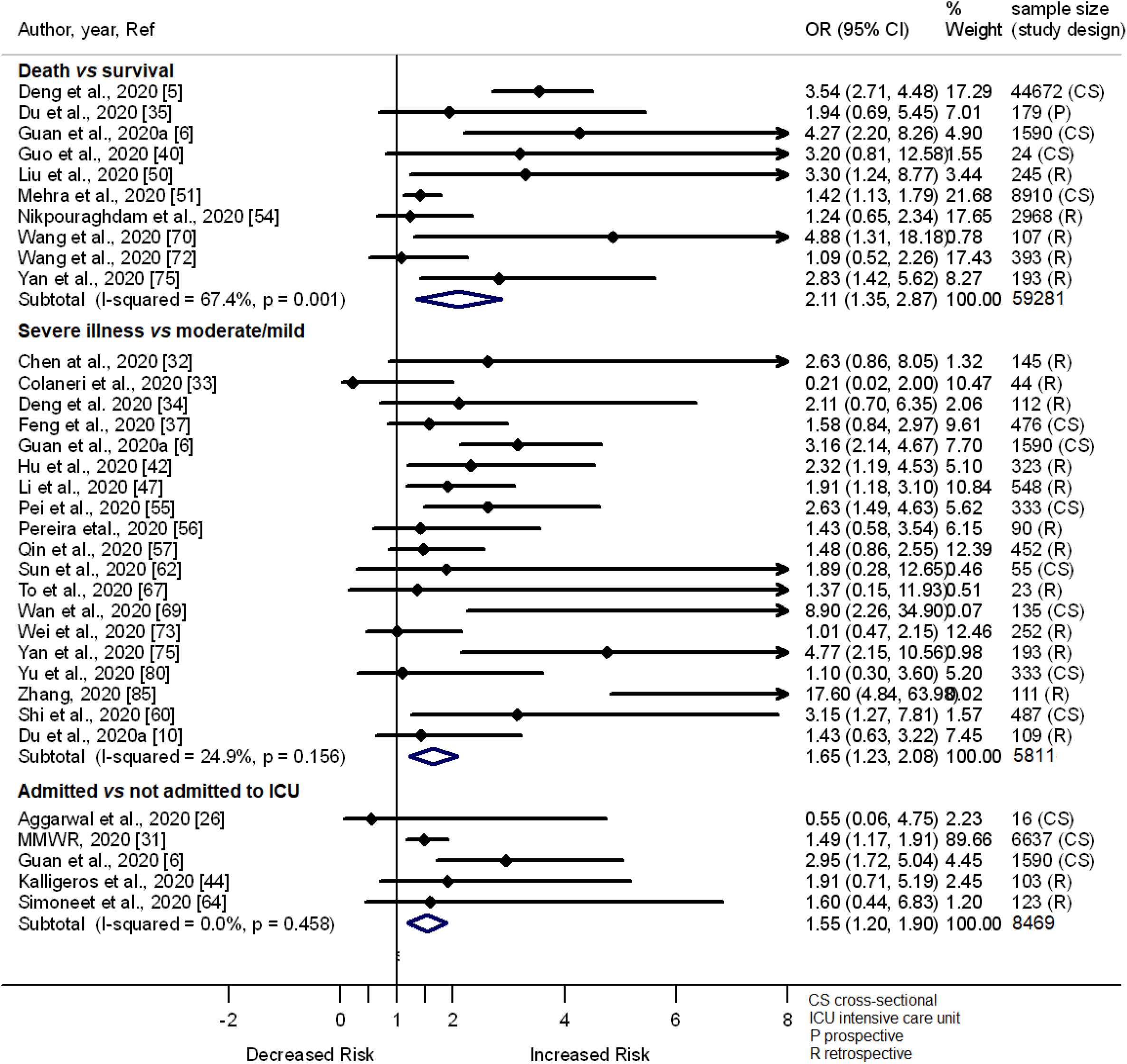
Forest plot showing the relationship of diabetes with the odds of death, severe illness and admission in ICU among COVID-19 patients.

### Chronic respiratory disease

Of 71 included studies, 19 compared the odds of death *vs* survival (n=9), severe *vs* moderate or mild illness (n=9), and admission to ICU *vs* not (n=4) among COVID-19 patients who had chronic respiratory disease *vs* those without this problem. Of the 9 studies which compared the odds of death *vs* survival, six reported significantly greater odds of death among COVID-19 patients with chronic respiratory disease, but three documented lack of association between chronic respiratory disease and the odds of death. On the other hand, out of the 9 studies which compared the odds of severe *vs* mild or moderate illness among COVID-19 patients, only three reported significantly greater odds of severe illness, but five showed a lack of association between chronic respiratory disease and the odds of developing severe illness. One study reported lower odds of severe illness among COVID-19 patients with chronic respiratory disease as compared to those without chronic respiratory disease. Meta-analysis of the 19 studies showed association of chronic respiratory disease with increased odds of death (OR 2.83, 95% CI 2.14– 3.51, I^2^= 0.0%, n=9, N=59,624) and admission to ICU (OR 1.52, 95% CI 1.09–1.94, I^2^ = 0.0%, n=5, N=8,346), but there was lack of correlation of the disease with severe illness (OR 1.39, 95% CI 0.15–2.64, I^2^ = 27.0%, n=9, N=48,381) among COVID-19 patients (Fig 5).

**Fig 5.**
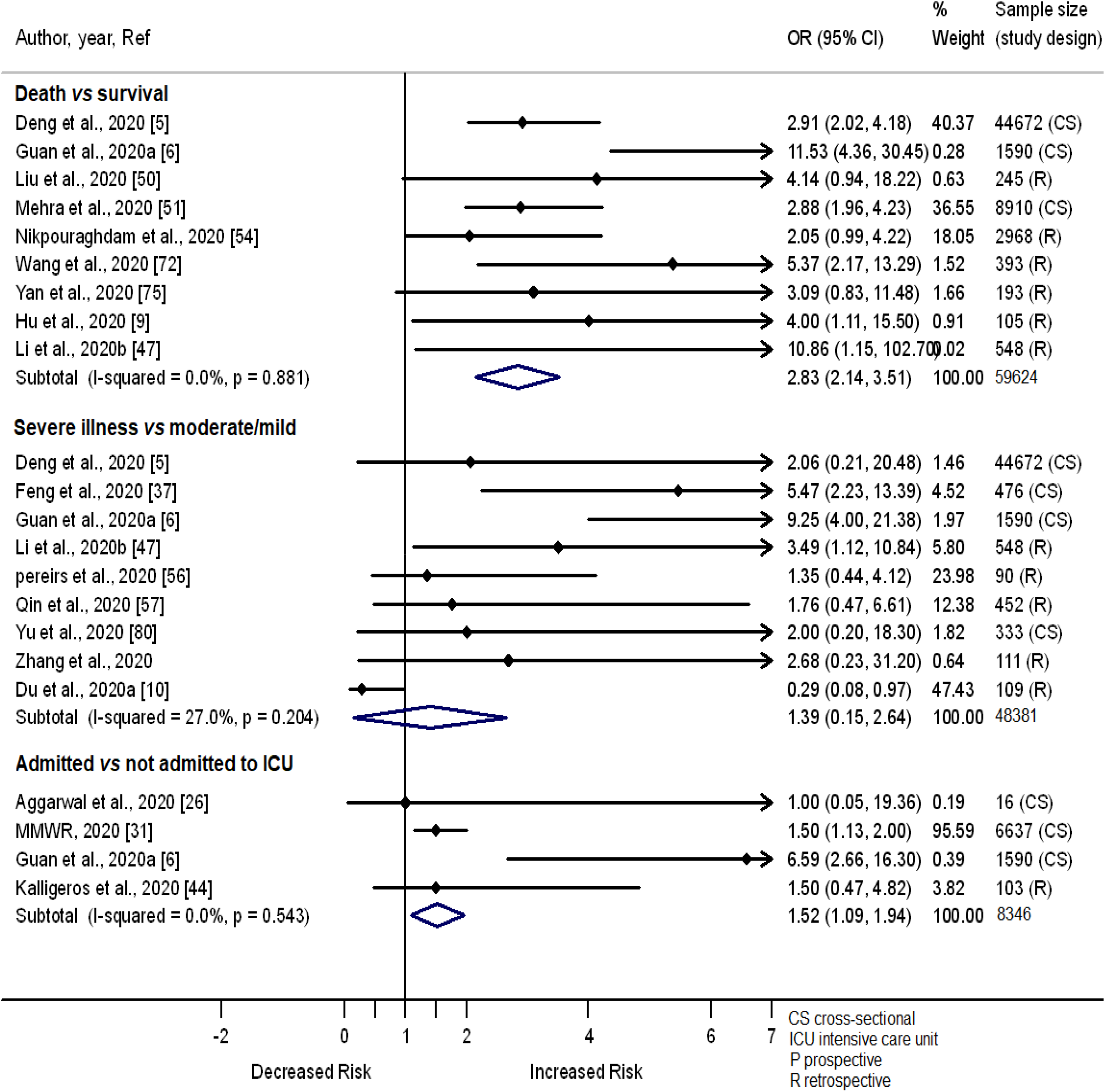
Forest plot showing the relationship of chronic respiratory diseases with the odds of death, severe illness and admission in ICU among COVID-19 patients.

### Cancer

Thirteen studies examined the nature of the relationship of cancer with the odds of death (n=5), severe illness (n=6) and admission to ICU (n=3) among COVID-19 patients. Increased odds of death, severe illness and admission to ICU among COVID-19 patients who had cancer was reported in 2 (out of 5), 3 (out of 6) and 1 (out of 3) studies, respectively. The remaining studies showed a lack of association between having cancer and the odds of death (n=3), severe illness (n=3) and admission to ICU (n=2) among COVID-19 patients. Meta-analysis of the 13 studies showed lack of association of cancer with the odds of death (OR 1.72, 95% CI 0.39 – 3.05, I^2^ = 0.0%, n=5, N=46,793), severe illness (OR 2.32, 95% CI 0.45 – 4.19, I^2^ = 0.0%, n=6, N=3,169) and admission to ICU (OR 1.79, 95% CI −0.74 – 4.31, I^2^ = 77.1%, n=3, N=1,804) among COVID-19 patients (S1 Fig).

### Chronic kidney disease

Twelve studies examined the nature of the relationship of chronic kidney disease with the odds of death (n=3), severe illness (n=7) and admission to ICU (n=2) among COVID-19 patients. Of the 12 studies, two showed increased odds of death (n=1) or severe illness (n=1) among COVID-19 patients who had *vs* didn’t have chronic kidney disease. The remaining 10 studies showed a lack of association of chronic kidney disease with the odds of death (n=3), severe illness (n=3) and admission to ICU (n=2). Meta-analysis of the 12 studies showed lack of association of chronic kidney disease with the odds of death (OR 2.32, 95% CI 0.06 – 4.58, I^2^ = 0.0%, n=3, N=4951), severe illness (OR 1.42, 95% CI 0.46 –2.38, I^2^ = 0.0%, n=7, N=7788) and admission to ICU (OR 1.37, 95% CI 0.88–1.86, I^2^ = 0.0%, n=2, N=6653) among COVID-19 patients (S2

Fig).

### Chronic liver disease

Out of the 71 studies included in this review, only seven tested the association of chronic liver disease with death or severe illness in COVID-19 patients. Of the seven studies, the only one reported a correlation of chronic liver disease with COVID-19 severity. A summary analysis of the seven studies showed lack of correlation of chronic liver disease with death (OR 1.41, 95% CI −2.20 – 5.02, I^2^ = 0.0%, n=2, N=352), severe illness (OR 1.35, 95% CI 0.67–2.03, I^2^ = 0.0%, n=4, N=6855) and admission to ICU (OR 1.77 95% CI 0.65 – 4.80, n=1, N=6637) (S3 Fig).

### Cerebrovascular diseases

Six studies reported findings on the odds of death (n=2), severe illness (n=2) and admission to ICU (n=2) among COVID-19 patients with cerebrovascular diseases *vs* those without having this comorbidity. Two studies by Guan et al. 2020 [6] and Wang et al. 2020 [72] reported increased odds of death among COVID-19 patients with cerebrovascular diseases. Another two studies also showed association of cerebrovascular diseases with increased odds of developing severe illness [37] and admission to ICU [8].

A meta-analysis of the two studies showed increased odds of death among COVID-19 patients with cerebrovascular diseases (OR 5.14, 95% CI 1.08 – 9.19, I^2^ = 0.0%, n=2, N=1,983), but the odds of severe illness (OR 2.18, 95% CI −0.07 – 4.44, I^2^ = 0.0%, n=2, N=928) and admission to ICU (OR 4.09, 95% CI −0.20 – 8.38, I^2^ = 0.0%, n=2, N=1,606) were similar between patients with cerebrovascular diseases and those without this comorbidity (S4 Fig).

### Smoking

A total of 14 studies included in the current meta-analyses that examined the impact of tobacco smoking on the clinical outcomes of COVID-19. Of these 14 studies, most showed a lack of association of smoking with death (2 out of 2), severe illness (9 out of 11), or admission to ICU (1 out of 1) among COVID-19 patients. Only two studies showed increased odds of severe illness among COVID-19 patients who were former or current smokers than non-smokers. A meta-analysis of the studies showed similar odds of death (OR 0.90, 95% CI 0.70 – 1.10, I^2^ = 0.0%, n=2, N=9155) or severe illness (OR 1.31, 95% CI 0.96 – 1.67, I^2^= 0.0%, n=11, N=4509) among COVID-19 patients who were former or current smokers than non-smokers (S5 Fig).

### Age

Out of the 71 studies, 27 examined the effect of age on the odds of death, severe illness and admission to ICU. Out of 27 studies, 15 treated age as >65 *vs* ≤ 65 years and 10 as ≥ 60 *vs* <60 years. Of the 15 studies which treated age as > 65 *vs* ≤ 65 years, 13 reported increased odds of death (n=5) or severe illness (n=9) among patients of ages > 65 years as compared to those of ≤ 65 years. Similarly, nine studies that treated age as ≥ 60 *vs* < 60 years reported increased odds of death (n=6) or severe illness (n=3) among patients of ages ≥ 60 years as compared to those < 60 years. Chen et al. [89] also showed an increased hazard ratio of severe illness among patients of Age ≥ 65 years as compared to younger individuals (Hazard ratio 3.43; 95% CI, 1.24-9.5). Dong et al. [88] also reported an increased prevalence of severe COVID-19 illness in children than those of adult patients. However, a study showed similar odds of severe illness among individuals of ages ≥ 60 *vs* <60 years [80]. Two studies also reported similar odds of admission to ICU among individuals of ages > 65 years *vs* those of ≤ 65 years [10,25]. A meta-analysis of these studies showed a greater odds of death (OR 3.56, 95% CI 1.21 – 5.90, I^2^ = 18.2%, n=6, N=13,964) and severe illness (OR 2.55, 95% CI 1.94 – 3.17, I^2^ = 24.5%, n=9, N=3374) among patients of ages > 65 years than those of ≤ 65 years. The summary odds ratio estimates of death (OR 6.09, 95% CI 3.53 – 8.66, I^2^ = 95.5%, n=6, N=100,733) and severe illness (OR 4.91, 95% CI 1.35 – 8.47, I^2^ = 0.0%, n=4, N=333) among older-aged COVID-19 patients than younger ones were even much greater when age was treated as ≥ 60 *vs* < 60 (Fig 6).

**Fig 6.**
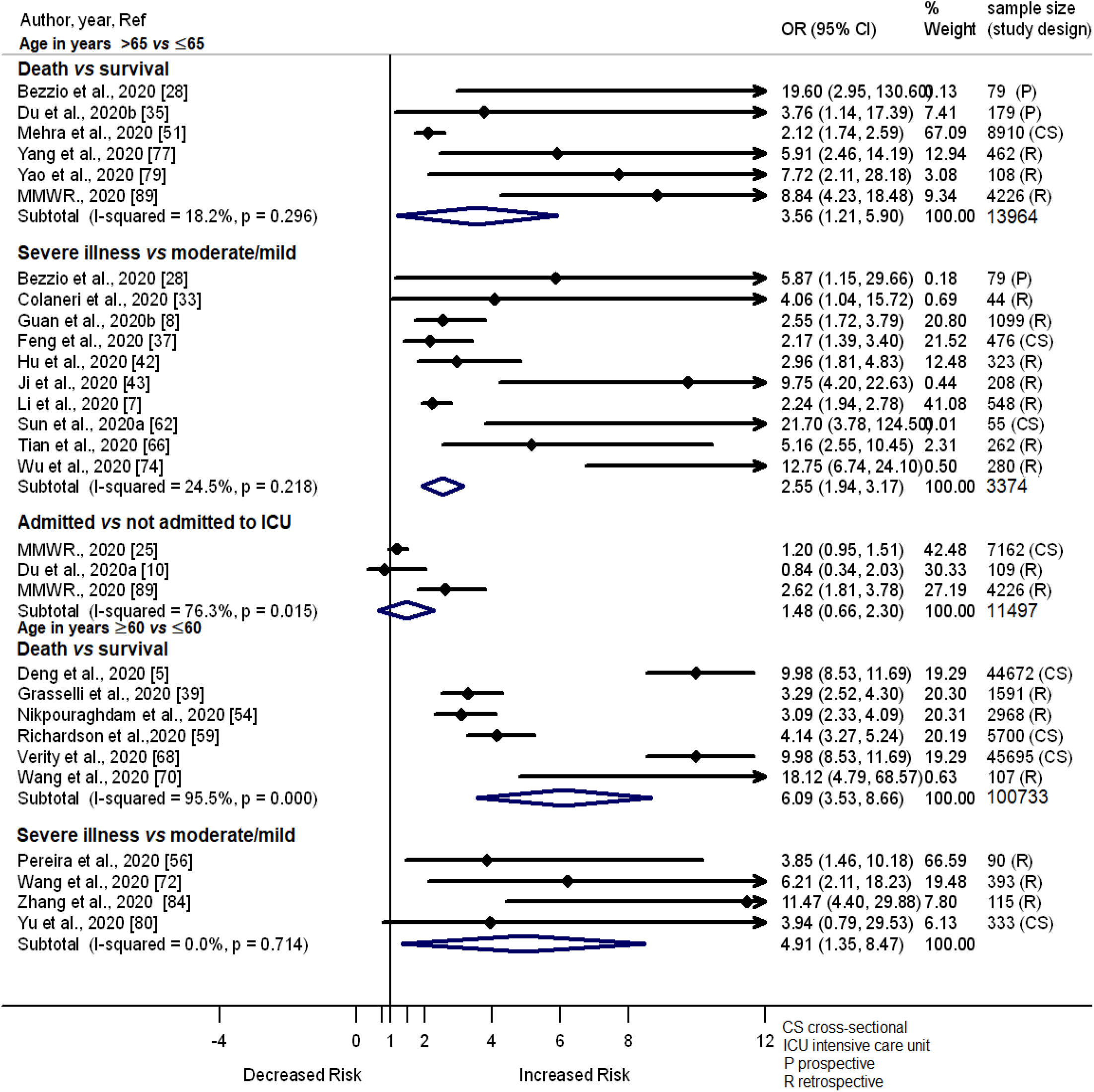
Forest plot showing the relationship of age with the odds of death, severe illness and admission in ICU among COVID-19 patients.

### Gender

Fifty-two studies examined the effect of gender on the odds of death (n=16), severe illness (n=32) and admission to ICU among COVID-19 patients (n=6). Out of 16 studies which compared the odds of death between males and females, 10 reported increased odds in males and six documented similar odds between males *vs* female. A meta-analysis of the 16 studies showed increased odds of death among males than females (OR 1.34, 95% CI 1.18 – 1.50, I^2^ = 38.7%, n=16, N=68,609). Similarly, of the 32 studies which compared the odds of severe illness between males *vs* females, 11 reported increased odds and 21 studies document similar odds between males *vs* female. A summary analysis of the 32 studies showed increased odds of severe illness among males than females (OR 1.35, 95% CI 1.23 – 1.47, I^2^ =0.0%, n=32, N=13,426); however, this could not be further assessed by tandem risk factors (e.g., underlying cardiovascular disease) in the available data.

There were six studies that examined the relationship of gender with the odds of admission to ICU. Out of these six studies, two showed increased odds of admission to ICU in males than females, but four studies showed a lack of association between gender and the odds of admission to ICU. A summary of the 6 studies showed a lack of correlation between gender and admission to ICU in COVID-19 patients (OR 1.52, 95% CI 0.69 – 2.36, I^2^ =2.9%, n=6, N=666) (Fig 7).

**Fig 7.**
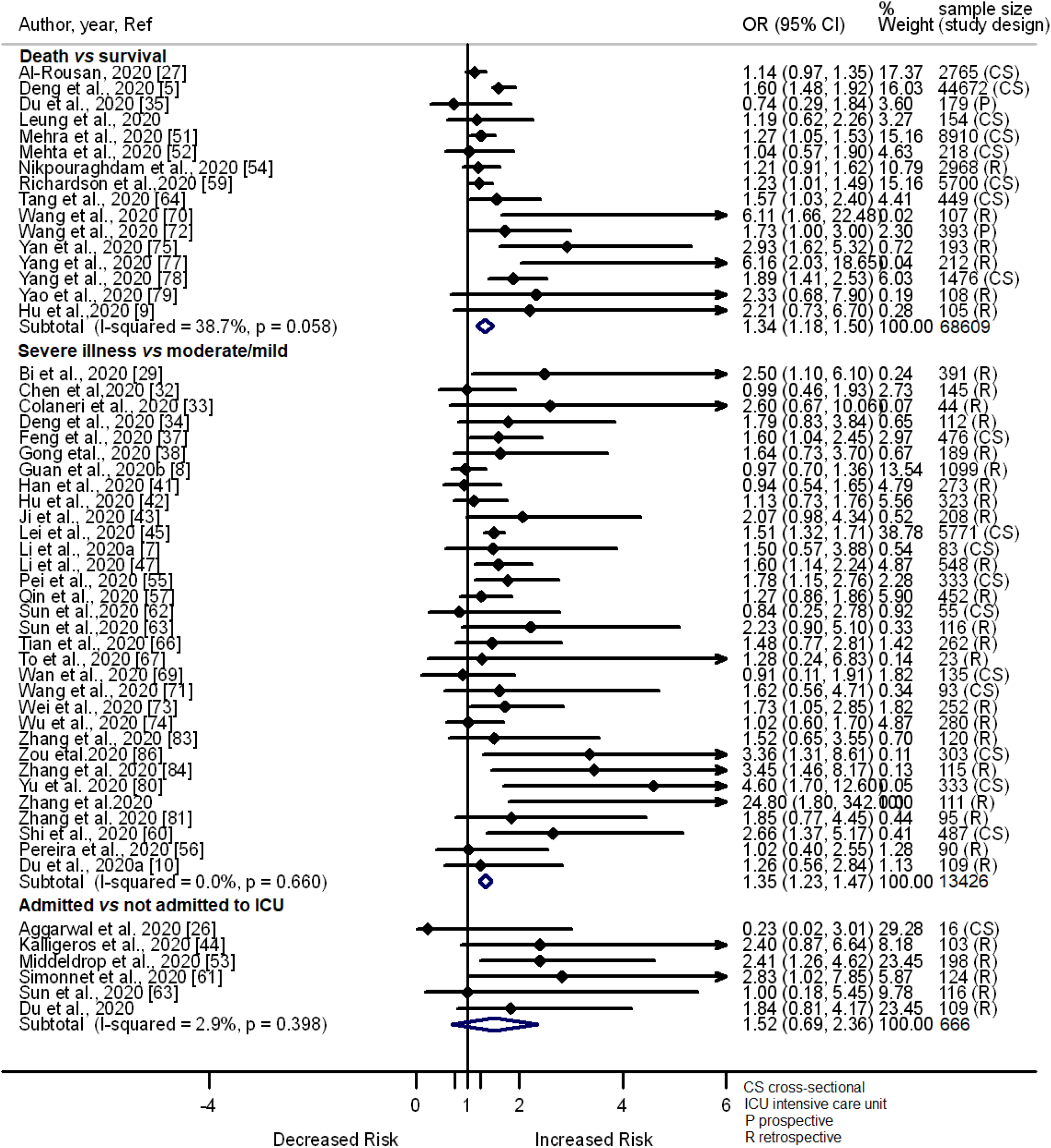
Forest plot showing the relationship of gender with the odds of death, severe illness and admission in ICU among COVID-19 patients.

### Heterogeneity assessment

There was no heterogeneity (I2= 0.0%) observed among the studies included in the meta-analyses that estimated the summary odds ratio of death, severe illness or admission to ICU among COVID-19 patients with cerebrovascular diseases, chronic liver disease, chronic kidney disease, cancer or smoker vs those without these comorbidities. There was also no or low heterogeneity among the studies included in the meta-analyses that examined correlation of clinical outcomes in COVID-19 patients with cardiovascular disease (I2= 0.0% for death and admission to ICU, I2=30. 6% for severe illness), diabetes (I2= 0.0% for admission to ICU, I2=24.9% for severe illness), chronic respiratory disease (I2= 0.0% for death and admission to ICU, I2=27.0% for severe illness), age in years > 65 *vs* ≤65 (I2= 18.2% for death, I2=24.5% for severe illness), and gender (I2= 0.0% for severe illness, I2=2.9% for admission to ICU, I2=38.7% for death). The heterogeneity level in meta-analyses performed to examine associations of hypertension with the odds of death (I2= 52.6%) and severe illness (I2= 47.8%) among COVID-19 patients was moderate.

However, heterogeneity was high among the studies combined in the meta-analyses to examine the association of death with diabetes (I^2^ = 67.4%) and age, ≥ 60 vs < 60 years (I^2^ = 95.5%) in COVID-19 patients. Studies that were included in the meta-analyses to assess the nature of the relationship of admission to ICU with hypertension (I^2^ = 79.2%) also had high heterogeneity. Subgroup analyses by study design dropped heterogeneity: diabetes (I^2^ = 0.0% in retrospective), and age (I^2^ = 0.0% in retrospective; I^2^ =0.0% in cross-sectional). Due to the few numbers of studies that reported results on the effect of hypertension on the risk of admission to ICU, we didn’t perform subgroup analyses by study region/country or study design. Removal of a study by Guan et al. 2020a [6], however, significantly minimized the heterogeneity of the studies that examined the effect of hypertension on the risk of admission to ICU (I^2^ = 1.0%).

On the other hand, the meta regression showed lack of effect of study area or country (meta regression coefficient (β)= −0.50, *P*= 0.073), study design (β = −0.23, *P*= 0.400) and sample size (1.25e-06, *P*= 0.912) on the log ORs of death among COVID-19 patients with diabetes vs those without this problem. Similarly, study area or country (β= −0.30, *P*= 0.380), study design (β = β = −0.04, *P*= 0.863) and sample size (β = 8.74e-06, *P*= 0.631) didn’t significantly predict the log ORs of admission to ICU among COVID-19 patients of age ≥60 years vs <60 years in the meta regression model.

### Publication bias assessment

Odds ratio distributions against their standard error estimates and Egger tests for asymmetry did not indicate significant publication bias: hypertension (bias 6.22, *P*=0.11), diabetes (bias 1.43, *P*=0.634), cardiovascular disease (bias 1.64, *P*=0.71), chronic respiratory diseases (bias 6.47, *P*=0.13), chronic kidney disease (bias 88.99, *P*=0.11), and cancer (Egger’s regression test (bias) 0.26, *P*=0.953) (Fig 8 and S6 Fig). Funnel plots of odds ratios of the likelihood of developing severe *vs* moderate or mild illness among COVID-19 patients and their Egger tests also were not significant: hypertension (bias 5.05, *P*=0.21); diabetes (bias 3.29, *P*=0.353), chronic respiratory diseases (bias −8.36, *P*=0.12); chronic kidney disease (bias −0.57, *P*=0.807); and, cancer (bias 3.34, *P*=0.598) (Fig 8 and S6 Fig). Studies comparing the odds ratio of admission to ICU *vs* not admission to ICU among COVID-19 patients with cancer (bias −6.27, *P*=0.51), hypertension (bias −5.36, *P*=0.12), diabetes (bias 0.53, P=0.763), cardiovascular (bias 1.37, *P*=0.40), chronic respiratory diseases (bias 1.98, *P*=0.67) were also spread evenly on both sides of the average OR estimates, creating an approximately symmetrical funnel-shaped distribution (Fig 8 and S6 Fig). The odds ratio estimates of the studies which examined the relationship of chronic liver disease (bias 0.47, *P*=0.752), cerebrovascular diseases (bias −5.83, *P*=0.395) and smoking behavior (bias 0.98, *P*=0.217) with severe illness or death in COVID-19 patients also had approximately symmetrical funnel-shaped distribution and the Egger’s test for the asymmetry of the plots were not significant (S6 Fig). However, there was publication bias among the studies comparing the odds of i) death *vs* survival in patients with age > 65 *vs* ≤65 years (bias 11.29, *P*=0.062) and males *vs* females (bias 2.90, *P*=0.046); ii) severe *vs* mild or moderate illness in patients with age > 65 *vs* ≤65 years (bias 14.63, *P*=0.032), males *vs* females (bias 2.18, *P*=0.039) and those with cardiovascular disease *vs* without having this problem (bias 5.62, *P*=0.0.034), the odds ratio estimates scattered asymmetrically in the funnel plot (Fig 8 and S6 Fig).

**Fig 8.**
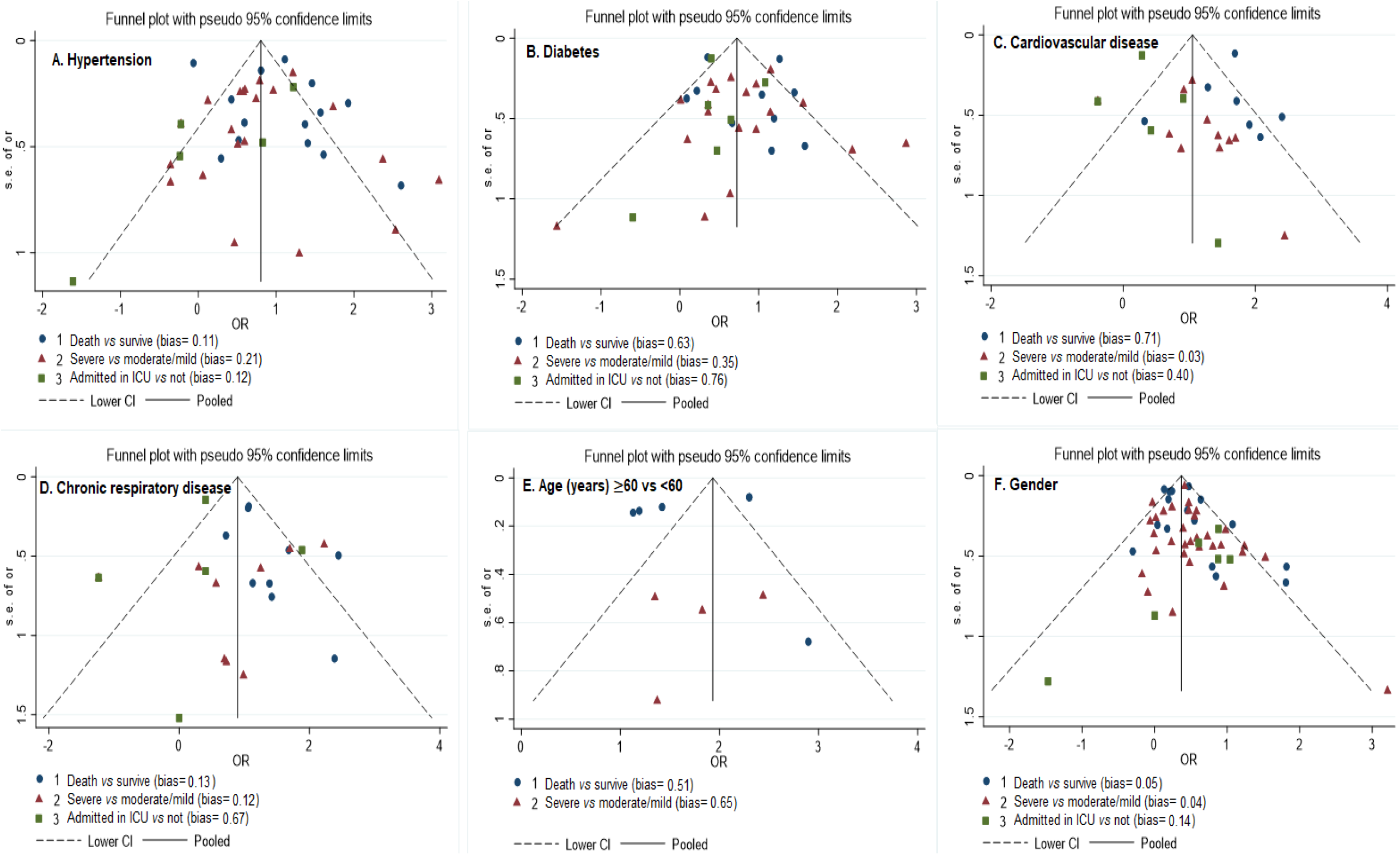
Funnel plots showing the odds ratio of death, severe illness and admission in ICU against the standard errors based on comorbidity and age.

### Risk of bias and quality of the studies

The study’s risk of bias and quality included in this study is summarized in the supplementary table (S4 table). Out of the 71 studies, 54 were moderate quality, and 18 were good quality in recruiting the study participants. The majority of the studies also used good (n=54) or moderate quality (n=17) reliable and valid tools to determine COVID-19 severity status and related deaths among the study participants. In many studies (n=44), the researchers or data collectors were not aware of the group (exposed vs. unexposed) to which the study participants belong in the data collection process and/or the participants were blinded about the research question. About 2/3 of the included studies were of moderate quality in terms of the study design (retrospective case-control or cohort) and controlling confounders that may affect the relationship between comorbidities and COVID-19 related outcomes or death. On the other hand, several studies rated as low quality based on study design (n=15), data collection (n=8), confounders (n=26), blinding (n=28) and dropouts and withdrawals (n=29). None of the studies were of low quality in terms of blinding of the outcome assessor and the study participants. Total rating using the six criteria showed that 14 studies were of strong quality, 23 studies were of moderate quality, and 34 studies were rated as low quality. Studies were included in this review, regardless of their qualities.

## Discussion

The current systematic review and meta-analysis confirmed the correlation of demographic factors and comorbidities with severe illness and death among COVID-19 patients, as reported in other reviews [11-13]. COVID-19 patients who were males, with age >60 or 65 years and those who had hypertension, diabetes, cardiovascular disease, chronic respiratory disease and cerebrovascular diseases, were found more susceptible to death. The risk of developing severe illness also increased among males COVID-19 patients of ages older than 60 or 65 years, and those who had hypertension, diabetes, and cardiovascular diseases. However, comorbidities, including cancer, chronic kidney and chronic liver diseases, were not associated with severe illness and mortality in COVID-19 patients.

Chronic diseases, including hypertension, diabetes, chronic respiratory disease, cardiovascular disease put the body in stress for a longer period of time. This stress can weaken the immune system. For example, diabetes can cause damage to β-cell through the release of inflammatory cytokines such as IL-1β and TNFα that [90]. Metabolic disorders may also impair macrophage and lymphocyte function leading to low immune function [39]. Chronic disease can also affect the function of different organs of the body such as the lung, heart, vascular tube. These organs dysfunction and low immunity due to the chronic diseases will complicate and increase the risk of severe illness and death during SARS-CoV-2 infection. It is also possible that angiotensin converting enzyme inhibitors or angiotensin receptor blockers used to treat diabetes, hypertension, or heart disease may upregulate Angiotensin converting enzyme 2, a receptor to SARS-CoV-2 in the host cell [91-93]. This may facilitate the viral multiplication within the host cell, though this is controversial [94].

Similarly, immunity can decrease with age, which will make older patients vulnerable to develop severe illness and death during SARS-CoV-2 infection. While functional immunosuppression of senescence may play a role in older adults experiencing worse disease, older adults also are more likely to have an underlying chronic disease. Males may also be more likely to involve in risky behaviors, including smoking, alcohol drinking and drug use, which can down-regulate the immune response against the SARS-CoV-2 infection.

Lack of association of some chronic diseases (e.g., kidney disease) and smoking with COVID-19 outcomes could be due to limitations in the original studies. The role of chronic kidney disease was assessed among comparatively few participants, which could explain its discordance with the results among cardiovascular equivalents. Potential ascertainment bias in the original studies could also partly explain the lack of association of chronic respiratory disease with severe illness though the disease showed to have a role in death and admission to ICU. Similarly, lack of correlation of smoking with clinical outcomes in the COVID-19 patients could be due the watering effect of the chronic disease outcomes. If most of the patients with cardiopulmonary diseases were smokers, exacerbated by ascertainment bias (they did not ask enough, or misinterpreted prior smoking and current smoking from never smoking), could miss the distinction of tobacco use. Unfortunately, most studies didn’t control chronic disease as a potential confounder when examining the role of smoking on COVID-19 related outcomes.

These findings have a number of public health and research implications that will help in the management of high-risk COVID-19 patients to mitigate the progression of the disease and associated death. Based on our findings, special strategies are warranted to prevent SARS-CoV-2 infection and manage COVID-19 cases with underlying comorbidities, particularly older age males’ patients. For example, creating awareness and provision of robust personal protection measures would be needed to prevent older aged individuals who have comorbidities from SARS-CoV-2 infection. Second, more intensive surveillance and early hospital referral would be important for older age males COVID-19 patients with comorbidities. When these high-risk older age COVID-19 patients, particularly those with comorbidities, are admitted to a hospital, special attention and treatment care could be considered to prevent further progression of the disease and death. Third, future vaccination intervention against SARS-CoV-2 infection should be formulated to better protect people with comorbidities. In addition, when the SARS-CoV-2 vaccination is available, people with hypertension, cardiovascular disease, diabetes, respiratory system disease and cerebrovascular disease should be a priority in the vaccination recommendations. The current findings also suggest prospective cohort studies with adequate power to verify whether and how comorbidities and demographics can affect the risk of acquisition of the SARS-CoV-2 and clinical outcomes of the disease.

This meta-analysis has a number of strengths. Unlike previous meta-analyses on the related topic [11-13], it i) involves a comprehensive analysis of a large number of studies based in China, South Korea, Iran, Europe and North America; ii) summarized the literature on the effect of comorbidities, including cancer, chronic liver and cerebrovascular disease with severe illness and death; and iii) compared the risk of admission to ICU vs not among hospitalized COVID-19 patients stratified based on comorbidities and demographic status. In addition, we found little publication bias regarding the examination of the association of comorbidities with severe illness and death in COVID-19 patients. Most of our meta-analyses had no or low heterogeneity, including the association of cardiovascular disease, chronic respiratory disease, chronic liver, cerebrovascular disease, chronic kidney disease, age, gender and severe illness and death. Although there was a high level of heterogeneity in studies that examined the correlation of diabetes with severe illness, subgroup analysis by the study design significantly decreased the heterogeneity. Variation in the study participant inclusion criteria and the methods used for examining comorbidities among the studies could have also contributed to the observed high heterogeneity.

However, the calculated summary OR in the current meta-analyses could have been affected due to limitations in the original studies. Most studies reported frequencies or crude estimates which were not adjusted for potential confounder that could affect the relationship between comorbidities and the risk of severe illness or death in COVID-19 patients. In addition, the sample size and/or the number of severe cases or deaths in patients with varied comorbidity or demographic status were small in some studies, increasing the confidence interval estimates for the OR and decreasing the power to reject false associations. These limitations in the original studies could have under or overestimated the summary OR estimates of the meta-analysis that examined the relationship of demographic and comorbidities with this review’s outcomes. Moreover, due to the cross-sectional and retrospective nature of the original studies, we cannot make firm conclusions that the observed higher odds of severe illness and death among COVID-19 patients were due to comorbidities, their demographic status, or other undetermined factors, though this was mitigated with the incorporation of random effects. In addition, as the majority of the studies included in the meta analysis were retrospective, we didn’t assess the risk of selection bias. Furthermore, in order to decrease the risk of misclassification bias, the exposures cardiovascular diseases, diabetes and cerebrovascular diseases could be merged while examining their effect on the risk of death and severe illness. However, the studies assessed and reported the results for these exposures differently, separately.

## Conclusions

In conclusion, older age and the presence of comorbidities or chronic diseases increase the risk of developing severe illness, admission to ICU and death among COVID-19 patients. Special strategies are warranted to prevent SARS-CoV-2 infection and manage COVID-19 cases with underlying comorbidities, particularly older age male patients, though the impact in women may be insufficiently observed. Prospective, longitudinal studies that control confounders are needed to assess strategies for mitigating severe illness, admission to ICU, and death in COVID-19 patients.

## Data Availability

All relevant data are within the manuscript and its supporting information files

## Supporting information

S1 Table. PRISMA 2009 Checklist.

S2 Table. Searching details Embase. S3 Table. Characterstics of the studies.

S4 Table. Risk of bias and quality of the studies included in this review.

S1 Fig. Forest plot showing the relationship of cancer with the odds of death, severe illness and admission in ICU among COVID-19 patients.

S2 Fig. Forest plot showing the relationship of chronic Kidney disease with the odds of death, severe illness and admission in ICU among COVID-19 patients.

S3 Fig. Forest plot showing the relationship of chronic liver disease with the odds of death, severe illness and admission in ICU among COVID-19 patients.

S4 Fig. Forest plot showing the relationship of Cerebrovascular disease with the odds of death, severe illness and admission in ICU among COVID-19 patients.

S5 Fig. Forest plot showing the relationship of smoking with the odds of death, severe illness and admission in ICU among COVID-19 patients.

S6 Fig. Funnel plots showing the odds ratio of death, severe illness and admission in ICU against the standard errors based on comorbidity and age status.

