## Supplemental Fig 1 for "Risk factors for severe illness and death in COVID-19: a systematic review and meta-analysis"

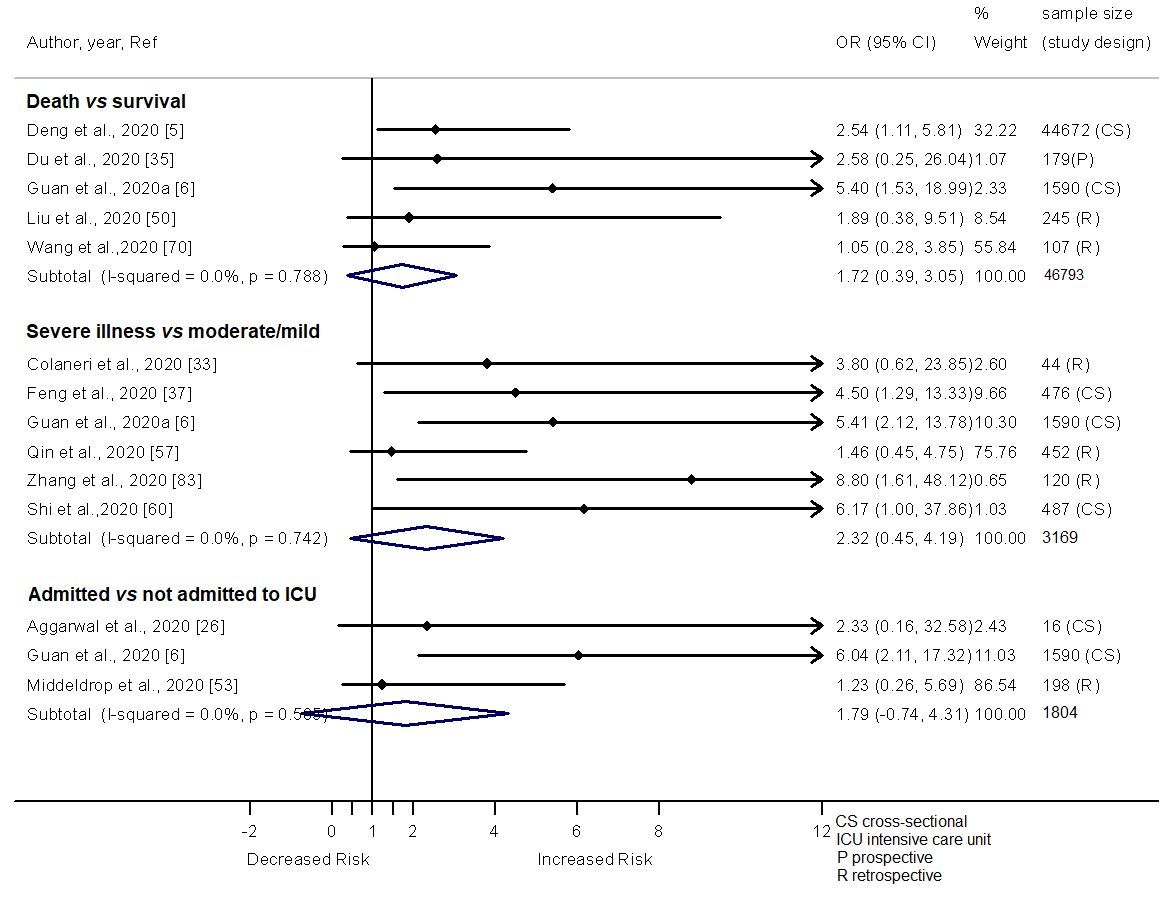
Additional file 4. Forest plot showing the relationship of cancer with the odds of death, severe illness and admission in ICU among COVID-19 patients.
