## Supplemental Fig 5 for "Risk factors for severe illness and death in COVID-19: a systematic review and meta-analysis"

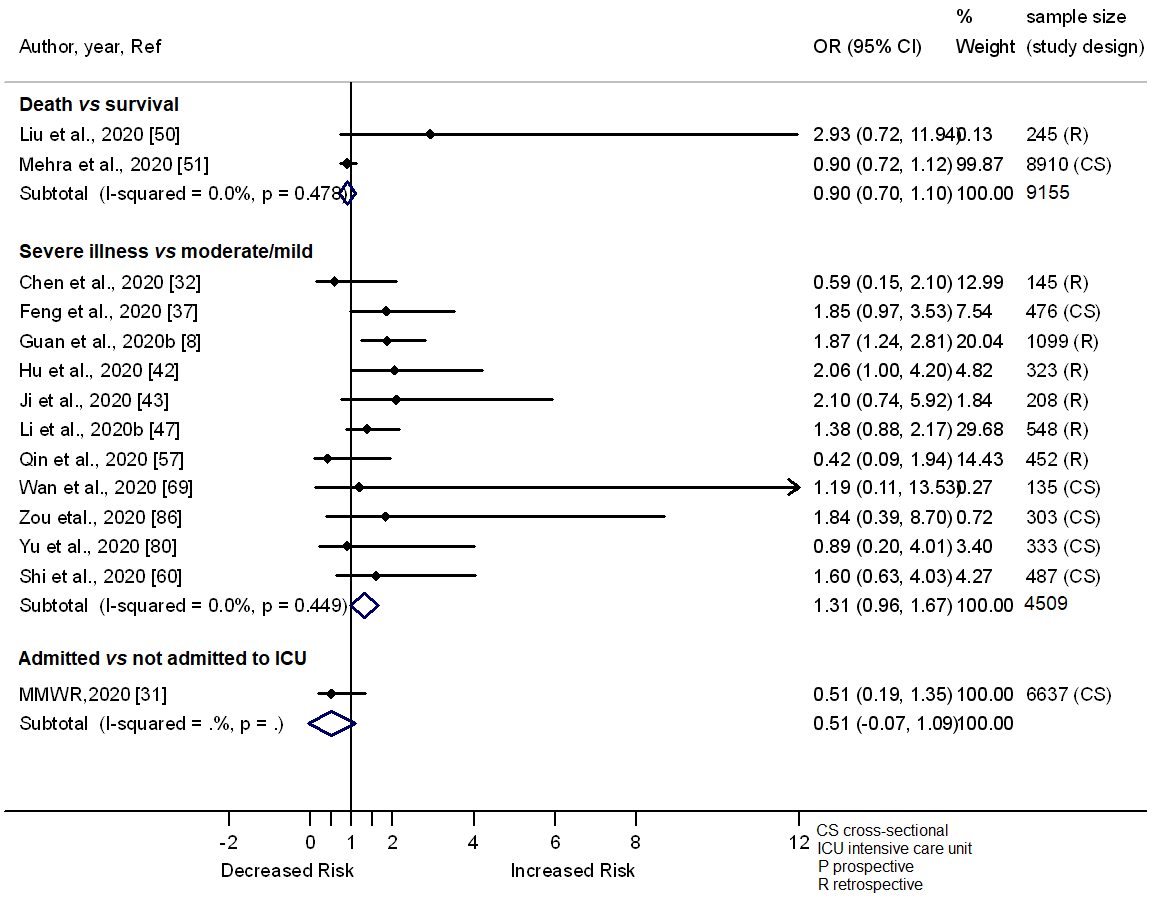

Additional file 8. Forest plot showing the relationship of smoking with the odds of death, severe illness and admission in ICU among COVID-19 patients.
