## Supplemental Fig 6 for "Risk factors for severe illness and death in COVID-19: a systematic review and meta-analysis"

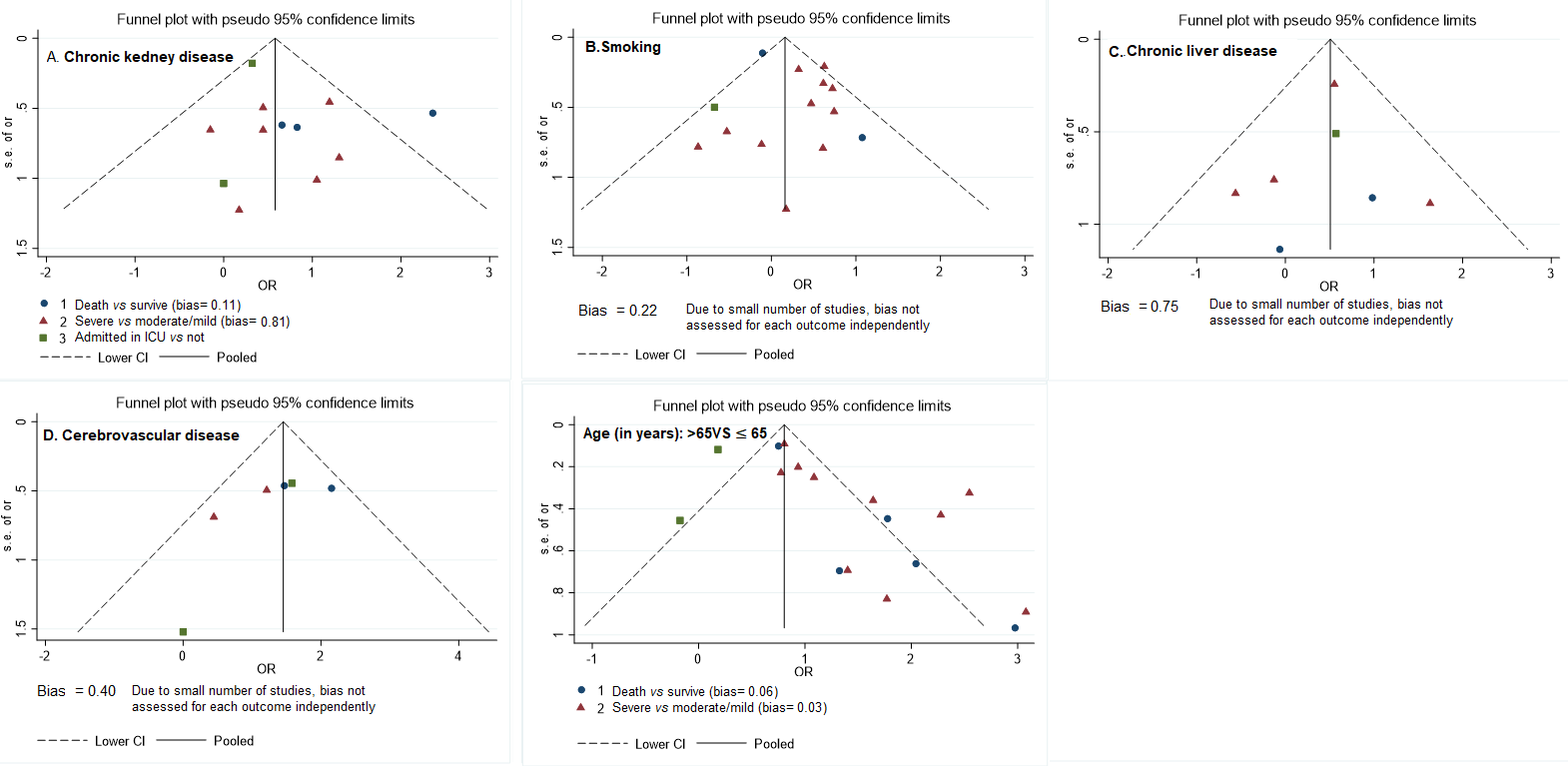
Additional file 9. Funnel plots showing the odds ratio of death, severe illness and admission in ICU against the standard errors based on comorbidity and age status
