## Supplemental Table 2 for "Risk factors for severe illness and death in COVID-19: a systematic review and meta-analysis"

Additional file 2. Search details Embase

('obesity'/exp OR obesity OR 'hypertension'/exp OR hypertension OR 'asthma'/exp OR asthma OR 'nutrition'/exp OR nutrition OR 'age'/exp OR age OR 'gender'/exp OR gender OR 'ethnicity'/exp OR ethnicity OR 'race'/exp OR race OR 'income'/exp OR income OR 'poverty'/exp OR poverty OR 'pregnancy'/exp OR pregnancy OR 'breast feeding'/exp OR 'breast feeding' OR 'medical conditions' OR medications OR 'chronic diseases'/exp OR 'chronic diseases' OR 'influenza'/exp OR influenza OR 'stroke'/exp OR stroke OR 'hiv'/exp OR hiv OR 'cancer'/exp OR cancer OR 'diabetes'/exp OR diabetes OR 'cardiovascular disease'/exp OR 'cardiovascular disease' OR 'coronary heart disease'/exp OR 'coronary heart disease' OR 'chronic respiratory disease'/exp OR 'chronic respiratory disease' OR 'sequential organ failure assessment'/exp OR 'sequential organ failure assessment' OR 'smoking'/exp OR smoking OR 'co infection'/exp OR 'co infection' OR 'comorbidity'/exp OR comorbidity OR comorbidities OR 'risk'/exp OR risk) AND ('clinical'/exp OR clinical OR severe OR 'complications'/exp OR complications OR 'mortality'/exp OR mortality OR 'death'/exp OR death) AND ('coronavirus disease 2019'/exp OR 'coronavirus disease 2019' OR 'covid-19'/exp OR 'covid-19' OR 'severe acute respiratory syndrome corona virus 2' OR 'sars cov 2'/exp OR 'sars cov 2' OR 'corona virus 2' OR 'sars corona virus')
