## Supplemental Table 3 for "Risk factors for severe illness and death in COVID-19: a systematic review and meta-analysis"

Additional file 3. Characteristics of the studies

|  |  |  |  | **Outcomes ( 95% CI)** | | |
| --- | --- | --- | --- | --- | --- | --- |
| **References ((Study area)** | **Age in years** | **Sample size** | **Study design** | **Death *vs* survival** | **Severe illness *vs* mild/moderate** | **Admission in ICU *vs* not in ICU** |
| Aggarwal et al. 2020 ((USA) [26] | 38-95 mean=65.5 median=67 | 16 | CS |  |  | - Male 0.23, 0.02 – 3.01 - Obese 2.77, 0.37 – 21.02 - Hypertension 0.2, 0.02 – 1.71 - Diabetic 0.55, 0.06 – 4.75 - Coronary artery 2.33, 0.16-32.58 - Cerebrovascular 1, 0.05 – 19.36 - Congestive cardiac failure - 4.2, 0.33 – 53.12 - Chronic kidney 5, 0.58-42.79 - COPD 1, 0.05 – 19.36 - Cancer 2.33, 0.16 – 32.58 |
| Al-Rousan, 2020 (South Korea) [27] | Not provided | 2765 | CS | - Males 1.14, 0.97 – 1.35 |  |  |
| Bezzio et al. 2020 (Italy)  [28] |  | 79 | P | - Age>65 19.6, 2.95 – 130.6 - Charlson comorbidity index (CCI) >1 16.66, 1.80 – 153.9 | - Age>65 5.87, 1.15 – 29.66 - Carlson comorbidity index CCI>1 2.91, 1.06 – 9.21 |  |
| Bi et al. 2020 (china) [29] | Mean 45 | 391 | R |  | - Male 2.5, 1.1-6.1 |  |
| Bi et al. 2020 (China) [30] | Median 46 | 113 | R |  | - Age>50 5.42, 1.92 15.26 - Male 1.13, 0.44-2.91 - Smoker 8.6, 1.88-39.55 - Comorbidity 2.21, 0.86-5.68 |  |
| MMWR, 2020  (APRIL3, USA) [31] | All ages | 6637 | CS |  |  | - Diabetes 1.49, 1.17-1.91 - Chronic lung disease   1.50,1.13-2.00   - Cardiovascular disease 1.33,1.04-1.71 - Immunosuppression 1.52, 1.01-2.29 - Chronic renal disease 1.38,0.97-1.96 - Pregnancy 0.28, 0.10-0.81 - Chronic liver disease 1.77, 0.65-4.80 - Other comorbidities 1.11, 0.88-1.40 - Smoking 0.51, 0.19-1.35 |
| MMWR,2020  (APRIL14, USA) [24] | All ages | 8945 | CS | - Age>65 9.94, 4.53-21.8 |  | - Age>65 1.81, 1.14-2.84 |
| Chen et al. 2020 (china)  [32] | Mean=47.5  Sd= 14.6 | 145 | R |  | - Continuous age   - Median severely ill (52.8 year)   - Median non-severely ill (45.3)   - P value=0.0001 - Males 0.99, 0.46-1.93 - Hypertension, 1.81, 0.71-4.62 - Diabetes 2.63, 0.86-8.05 - Chronic liver disease 5.12,   0.90-29.14   - Chronic kidney disease   1.19, 0.11-13.48   - Smoking 0.59, 0.15-2.10 |  |
| Colaneri et al. 2020 ((Italy) [33] | Median=67.5  Range 10-94 | 44 | R |  | - Male 2.6, 0.67-10.06 - Age>65 4.06, 1.04-15.72 - Other comorbidities 2.6, 0.67-10.06 - Cancer 3.8, 0.62-23.85 - Heart disease 2.4, 0.59-9.63 - Hypertension 0.70, 0.19-2.61 - Diabetes 0.21, 0.02-2.00 |  |
| Deng G et al., 2020 (IChina) [5] |  | 44,672 | CS | - Males 1.6, 1.48-1.92 - Age>=60 9.98, 8.53-11.69 - Hypertension 2.87,2.39-3.45 - Diabetes 3.54, 2.71-4.48 - Cardiovascular disease 5.00 3.96-6.31 - Respiratory disease 2.58, 1.78-3.72 - Cancer 2.23, 0.97-5.11 |  |  |
| Deng et al, 2020  (China) [34] | Median 65  Range 24-92 | 112 | R |  | - Continuous age   - Severe median=68   - Non-severe median=56   - P value <0.01 - Male1.79 0.83-3.84 - COPD 2.06 0.21-20.48 - Hypertension1.53, 0.67-3.51 - Diabetes 2.11, 0.70-6.35 - Cardiovascular disease   1.73,0.51-5.81 |  |
| Du et al. 2020b [35]  (China) | Mean 57.6 ± 13.7  Range 18-87 | 179 | P | - Age >=65 3.76, 1.14-17.39 - Cardiovascular disease 2.46, 0.75 – 8.04 - Hypertension 4.08,1.58-10.5 - Male 0.74, 0.29-1.84 - Diabetes 1.94, 0.69-5.45 - Cancer 2.58, 0.25-26.04 |  |  |
| Du et al., 2020a  (China) [10] | Mean 70.7 | 109 | R |  | - Age >=65 0.84, 0.34-2.03 - Male 1.26, 0.56, 2.84 - Hypertension 0.80, 0.37-1.73 - Cardiovascular disease   0.68, 0.30-1.52   - Diabetes1.43, 0.63-3.22 - Chronic respiratory disease   0.29, 0.08-0.97 |  |
| Dudley et al. 2020  (China, South Korea) [36] |  | 52427 | CS | - Sex specific case fatality rate in china   - Males 2.8%   - Females 1.7% - Sex specific case fatality rate in Korea   - Males 1.19%   - Female 0.52% |  |  |
| Feng et al. 2020  (China) [37] | Median 53  IQR 40-64 | 476 | CS |  | - Age >=65 2.17,1.39-3.40 - Male1.60, 1.04 – 2.45 - Smoker 1.85, 0.97-3.53 - Other comorbidity 2.18, 1.37-3.47 - Hypertension1.82, 1.15-2.87 - Cardiovascular disease 2.50,1.27-4.92 - Diabetes 1.58, 0.84-2.97 - Cancer 4.15, 1.29-13.33 - Cerebrovascular disease 3.36,1.28-8.92 - Immunosuppression 7.35, 1.40-39.40 - COPD 5.47, 2.23-13.39 - Chronic renal 2.86, 0.39-20.58 |  |
| Gong et al., 2020  (china) [38] | Median 49 | 189 | R |  | - Median age for severe 63.5 - Median age for non-severe 45   - P value <0.01 - Male 1.64, 0.73-3.70 |  |
| Grasselli et al.2020  (Italy) [39] | Median 63  IQR 56-70 | 1591 | R | - Age >60 3.29, 2.52-4.30 - Hypertension 2.24,1.70-2.95 |  |  |
| Guan et al., 2020a  (China) [6] | Mean 48.9 | 1590 | CS | - COPD 11.53, 4.36-30.45 - Diabetes 4.27, 2.20-8.26 - Hypertension 6.86, 3.85-12.19 - Cardiovascular disease 5.56, 2.48-12.45 - Cerebrovascular disease 8.61, 3.35-22.12 - Cancer 5.40, 1.53-18.99 - Chronic renal disease10.58, 3.71-30.15 | - COPD 9.25, 4.00-21.38 - Diabetes 3.16, 2.14-4.67 - Hypertension 3.38, 2.50-4.57 - Cardiovascular disease   2.84,1.62-4.96   - Cerebrovascular disease   5.52, 2.66-11.45   - Cancer 5.41, 2.12-13.78 - Chronic renal disease3.30, 1.35-8.06, - Immunodeficiency 2.630.23-29.18 | - COPD 6.59, 2.66-16.30 - Diabetes 2.95, 1.72-5.04 - Hypertension 3.39, 2.21-5.21 - Cardiovascular disease   2.48, 1.14-5.38   - Cerebrovascular disease   4.85, 2.03-11.61   - Cancer 6.04, 2.11-17.32 |
| Guan et al, 2020b  (China) [8] | Median 47 | 1099 | R |  | - Age >=65 2.55, 1.72-3.79 - Male 0.97, 0.70-1.36 - Smoking 1.87, 1.24-2.81 |  |
| Guo et al., 2020  (China) [40] | Median 59 | 24 | R | - Diabetes 3.2, 0.81-12.58 |  |  |
| Han et al., 2020  (China) [41] | Median:  58.95 ±10.80  Severe cases:  58.97 ± 14.38  Critical Cases: 57.27 ± 17.25 | 273 | R |  | - Male 0.94, 0.54-1.65 |  |
| Hu et al., 2020  (China) [42] | Median 61 | 323 | R |  | - Age>=65 2.96, 1.81-4.83 - Male 1.13, 0.73-1.76 - Smoking 2.06, 1.0-4.2 - Hypertension 1.78, 1.11-2.88 - Diabetes 2.32, 1.19-4.53 |  |
| Ji et al., 2020 (China) [43] | Mean 44 ±16.3 | 208 | R |  | - Age   - Severe mean 57.7   - Non-severe mean 40.7   - P value <0.001 - Age (category)>65 9.75, 4.20-22.63 - Male 2.07, 0.98-4.34 - Smoking 2.10, 0.74-5.92 |  |
| Kalligeros et al., 2020  (China) [44] | Median 60 | 103 | R |  |  | - Continuous age 1.03, 1.0-1.07 - Non-Hispanic black 0.80, 0.26-2.45 - Male 2.40, 0.87-6.64 - Obesity 5.39,1.13-25.64 - Diabetes 1.91, 0.71-5.19 - Hypertension 0.79, 0.27-2.28 - Lung disease 1.50, 0.47-4.82 - Cardiovascular disease   1.52, 0.47-4.82 |
| Lei et al., 2020  (China) [45] | Median 56 | 5771 | CS |  | - Age continuous   - Severe median 59   - Non-severe median 55   - P value <0.001 - Male 0.67, 0.59-0.76 - Chronic liver disease 1.74, 1.08-2.80 |  |
| Leung et al., 20202  (China) [46] |  | 154 | CS | - Age continuous 1.04, 1.00-1.10 - Male 1.19, 0.62-2.26 |  |  |
| Li et al., 2020 (China) [47] |  | 83 | CS |  | - Age continuous - Severe mean age 25 - Non severe mean age 41.9   - P value <0.001 - Male 1.5, 0.57-3.88 - Hypertension 1.59, 0.24-10.18 |  |
| Li et al., 2020  (China) [7] | Median 60  IQR 48-69 | 548 | R |  | - Age>=65 2.24, 1.94-2.78 - Male 1.60, 1.14-2.24 - Smoking1.38, 0.88-2.17 - Chronic lung disease 3.49, 1.12-10.84 - Diabetes 1.91,1.18-3.1 - Hypertension 2.20, 1.51-3.20 - Chronic kidney disease   1.56, 0.43-5.60 |  |
| Lian et al., 2020 (China) [48] |  | 788 | R |  | - Age>=60 4.60, 2.80-7.55 | - Age>=60 7.55, 3.15-18.05 |
| Liu et al., 2020 (China) [50] | Mean 53.95 (16.90) | 245 | R | - Age continuous 1.09, 1.06-1.13 - chronic lung disease 4.14, 0.94-18.22 - Hypertension 3.94, 1.82-8.53 - Diabetes 3.30, 1.24-8.77 - Cancer 1.89, 0.38-9.51 - Chronic liver disease 2.67, 0.50-14.37 - Smoking 2.93, 0.72-11.94 |  |  |
| Mehra et al., 2020  (USA) [51] | Mean 49±16 | 8910 | CS | - Age>65 2.12, 1.74-2.59 - Males 1.27, 1.05-1.53 - Diabetes 1.42, 1.13-1.79 - Hypertension 0.94, 0.76-1.15 - COPD 2.88, 1.96-4.23 - Smoking 1.17 , 0.95-1.44 - Immunosuppression 1.60, 1.02-2.5 |  |  |
| Mehta et al., 2020  (USA) [52] | Median 69 | 218 | CS | - Males 1.04, 0.57-1.90 - Age   - Survived median 66   - Died median 76 |  |  |
| Middeldrop et al., 2020  (Netherlands) [53] | Mean 61 ± 14 | 198 | R |  |  | - Age   - Mean for intensive care=62   - Mean for non-intensive= 60   - P value 0.28 - Male 2.41, 1.26-4.62 - Cancer 1.23, 0.26-5.69 |
| Nikpouraghdam et al.,2 2020 (Iran) [54] | Mean 55.5 ± 15.15 | 2968 | R | - Age>=60 3.95, 2.98-5.23 - Male1.45,1.08 - 1.96 - Diabetes 1.35, 0.71-2.55 - Chronic respiratory disease 2.23, 1.08-4.59 - Hypertension   1.97, 0.92-4.21   - Cardiovascular disease   1.51, 0.53-4.30   - Chronic renal disease 2.49, 0.71-8.69 |  |  |
| Pei et al., 2020  (China) [55] | Mean 56.3 | 333 | R |  | - Male 1.78, 1.15-2.76 - Hypertension 1.70, 1.05-2.73 - Diabetes 2.61, 1.48-4.59 |  |
| Pereira et al., 2020  (USA) [56] | Median 57 | 90 | R |  | - Age >60 1.02, 0.40-2.55 - Hispanic 0.68, 0.27-1.68 - Hypertension 1.66, 0.64-4.38 - Diabetes 1.43, 0.58-3.54 - Chronic Lung disease 1.35, 0.44-4.12 - Chronic kidney disease   1.56, 0.59-4.11 |  |
| MMWR, 2020 (March 28^th^ ,2020) [90] |  | 7,162 | CS |  |  | - Age>=65 1.20, 0.95-1.51 |
| Qin et al., 2020 (China) [57] | Median 58 | 452 | R |  | - Male1.27, 0.86-1.86 - Smoking 0.42, 0.09-1.94 - Chronic lung disease 1.76, 0.47-6.61 - Hypertension 2.62, 1.65-4.17 - Cardiovascular disease   4.97, 1.47-19.79   - Cerebrovascular disease 1.56, 0.40-5.97 - Chronic liver disease 0.57, 0.11-2.88 - Diabetes 1.48, 0.86-2.55 - Cancer 1.46, 0.45-4.75 - Chronic kidney disease   0.86, 0.24-3.12 |  |
| Qu et al., 2020  (China) [58] | Median 20.5 | 30 | CS |  | - Age continuous   - Median age severe=60   - Median age non-severe 49.44   - P value 0.041 |  |
| Richardson et al., 2020  (USA) [59] | Median age 63  IQR 52-75 | 5700 | CS | - Age >=60 4.14, 3.27-5.24 - Male 1.23, 1.01-1.49 |  |  |
| Simonnet et al., 2020  (France) [61] | Median 60  IQR 51-70 | 124 | R |  |  | - Age continuous1.00, 0.97-1.04 - Male 2.83, 1.02-7.85 - Diabetes 1.60, 0.44-6.83 - Hypertension 2.29, 0.89-5.84 |
| Sun et al., 2020  (China) [62] | Median 44  IQR 34-56 | 55 | CS |  | - Age >65 21.71, 3.78-124.54 - Male 0.84, 0.25-2.78 - Hypertension 12.6, 2.18-73.4 - Diabetes 1.89, 0.28-12.65 |  |
| Sun et al., 2020  (China)[63] | Median 50  IQR 41-57 | 116 | R |  | - Age continuous   - Median non-severe   - 47   - Median severe 62   - P value <0.001 - Male 2.23, 0.90-5.51 | - Age continuous   - Median non-severe 57   - Median 71   - P value 0.008 - Male 1.00, 0.18-5.45 |
| Tang et al., 2020  (China) [64] | Mean 65.1  ± 12.0 | 449 | CS | - Age continuous   - Mean survivors 63.7   - Mean non-survivors 68.7   - Pvalue= <0.001   - Male 1.57, 1.03-2.40 |  |  |
| Tedeschi et al., 2020  (Italia) [65] |  | 609 | R | - Hypertension 4.31, 2.91-6.39 |  |  |
| Tian et al., 2020 (China) [66] | Median 47.5 | 262 | R |  | - Age >=65 5.16, 2.55-10.45 - Male1.48, 0.77-2.81 |  |
| To et al., 2020  (China) [67] | Median age 62 | 23 | R |  | - Male1.28, 0.24-6.83 - Hypertension 3.66, 0.51-26.2 - Diabetes 1.37, 0.15-11.93 |  |
| Verity et al., 2020  (China) [68] |  | 45,695 | CS | - Age >=60 9.98, 8.53-11.69 |  |  |
| Wan et al., 2020 (China)[69] | Median 47 | 135 | CS |  | - Age continuous   - Median mild 44   - Median severe 56   - P value <0.0001 - Male 0.91, 0.43-1.91 - Smoking 1.19, 0.11-13.53 - Diabetes 8.90, 2.26-34.9 - Hypertension 1.06, 0.30-3.67 |  |
| Wang et al., 2020 (China)[70] | Median 51  IQR 36-65 | 107 | R | - Age >=60 18.12, 4.79-68.57 - Male 6.11, 1.66-22.48 - Hypertension 5, 1.74-14.30 - Cardiovascular disease 7.97, 2.29-27.75 - Diabetes 4.88, 1.31-18.18 - Chronic liver disease 0.94, 0.10-8.57 - Cerebrovascular diseases   5.31, 0.98-28.71 |  |  |
| Wang et al., 2020  (China) [71] | Mean 52.1  Sd 18.07 | 93 | CS |  | - Age >60 6.21, 2.11-18.23 - Male 1.62, 0.56-4.71 |  |
| Wang et al., 2020  (China) [72] | Mean 71+-8 | 393 | P | - Age continuous   - Survived median 68   - Died median 76   - P value <0.001 - Male 1.73, 1.00-3.00 - Hypertension 1.53, 0.89-2.64 - Diabetes 1.09, 0.52-2.26 - Cardiovascular disease   3.60, 1.90-6.82   - Cerebrovascular disease 4.34, 1.75-10.73 - Chronic kidney disease 1.93, 0.57-6.47 - COPD 5.37, 2.17-13.29 - Cancer 1.05, 0.28-3.85 |  |  |
| Wei et al, 2020  (China) [73] | Mean  64.8 ± 13.3 | 252 | R |  | - Male 1.73, 1.05-2.85 - Diabetes 1.01, 0.47-2.15 - Hypertension 2.10, 1.22-3.60 - Cardiovascular disease - 3.56, 1.25-10.13 |  |
| Wu et al.,2020 (China) [74] | Mean 43.12  SD 10.02 | 280 | R |  | - Age ≥65 12.75. 6.74-24.1 - Male 1.02, 0.60-1.70 |  |
| Yan et al., 2020  (China) [75] | Median 64 (IQR 49–73) | 193 | R | - Age continuous   - Survivor median 46   - Non-survivor median 70   - P value <0.01 - Diabetes 4.77, 2.15-10.56 - Male 2.93, 1.62-5.32 - Diabetes 2.83, 1.42-5. - Hypertension 4.81, 2.48-9.35 - Cardiovascular disease   6.75, 2.25-20.16   - Chronic respiratory disease 3.09, 0.83-11.48 | - Diabetes 4.77, 2.15-10.56 | - Diabetes 2.83, 1.42-5.62 |
| Yang G et al., 2020  (China) [76] |  | 462 | R | - Hypertension 1.68, 0.67-4.21 | - Hypertension 1.13, 0.65-1.98 |  |
| Yang R et al., 2020  (China) [77] | Median 55.6  IQR=40-67 | 212 | R | - Age ≥65 5.91, 2.46-14.19 - Male 6.16, 2.03-18.65 |  |  |
| Yang X et al., 2020  (China) [78] | Median 57  IQR 47-67 | 1476 | CS | - Continuous age   - Non survivors mean 67   - Survivors mean 56 - Male 1.89, 1.41-2.53 |  |  |
| Yao et al., 2020  (China) [79] | Median 52  IQR 37-58 | 108 | R | - Age>65 7.72, 2.11-28.18 - Male 2.33, 0.68-7.9 - Hypertension 13.53, 3.55-51.5 |  |  |
| Zhang R et al., 2020  (China) [83] | Mean 45.4  SD 15.6 | 120 | R |  | - Age continuous 1.1, 1.0-1.1 - Male 1.52, 0.65-3.55 - Hypertension 10.70, 3.56-32.12 - Cardiovascular disease 4.3, 1.07-17.2 - Malignancy 8.8, 1.61-48.12 |  |
| Zou et al.2020 (China)  [86] |  | 303 | CS |  | - Male 3.36, 1.31-8.61 - Age: Mild (50 years), severe (65), P= < 0.001 - Chronic underlying disease 4.8, 1.95- 11.82 - Smokers 1.84, 0.39-8.7 |  |
| Zhang et al. 2020 (China) [82] | 38-91 | 19 | R | - Hypertension 0.72, 0.11-4.62   - Diabetes 0.17, 0.01-2.04   - Cerebrovascular disease 0.3, 0.02-4.06 |  |  |
| Zhang et al. 2020 (China) [84] | 20-86 | 115 | R |  | - >=60 years 11.47, 4.40-29.88 - Male 3.45, 1.46-8.17 |  |
| Zhang et al.2020 (China) [85] | 32-81 | 111 | P |  | - Male 24.8, 1.8−342.1 - Comorbidities 52.6, 3.6−776.4 - Age as a continuous P<0.001 - Hypertensive 22, 6.03-80.3 - Cardiovascular disease   11.5, 0.98-134.4   - Chronic obstructive pulmonary disease: 2.68 CI: 0.23-31.20 - Diabetes 17.6, 4.84- 63.98 |  |
| Zhang et al. 2020 (China) [81] | 23-88 | 95 | R |  | - 40–60 years 0.54 CI: 0.23-1.28) - Male 1.85 CI: 0.77-4.46 |  |
| Yu et al. 2020 (China)  [80] | 0-88 years | 333 | CS |  | - <60 years10.79, 3.94- 29.53 - Male 4.6, 1.7-12.6) - Heart disease 4.2, 1.2-14.2 - Diabetes 1.1 CI: 0.3-3.6 - Hypertensive 0.7, 0.2-2.0 - Respiratory disease 2.0, 0.2-18.3 - Smokers 0.89, 0.20 -4.01 - Alcohol 0.62, 0.22-1.70 |  |
| Hu et al., 2020 (China) [9] | Mean 60  Sd 16.32 | 105 | R | - Male 2.21, 0.73-6.70 - Age continuous   - Mean non survivors=75   - Mean survivors=57   - P value<0.01 - Hypertension 1.34, 0.45-3.96 - Chronic pulmonary disease 4.0, 1.11-15.50 |  |  |
| Shi et al., 2020  (China) [60] |  | 487 | CS |  | - Age continuous   - Mean severe=56   - Mean non severe=45   - P value <0.001 - Male 2.66, 1.37-5.17 - Smoking 1.60, 0.63-4.03 - Hypertension 5.65, 3.05-10.45 - Diabetes 3.15, 1.27-7.81 - Cardiovascular disease   5.47, 1.54-19.41   - Cancer 6.17 1.00-37.86 - Chronic liver disease 0.88, 0.20-3.92 - Chronic renal disease 3.68, 0.69-19.52 |  |
| Chen R et al., 2020  (China) [87] | 65 to 74 years | 1590 | R |  | - Age≥65 (HR*, 3.43; 95% CI, 1.24-9.5), - Coronary heart disease (HR* 4.28; 95% CI, 1.14-16.13) - Cerebrovascular disease (HR* 3.1; 95% CI, 1.07-8.94) |  |
| Dong Y et al., 2020  (China) [88] | Median 7 years | 2135 | R |  | - Children showed less severe COVID-19 illness than those of adult patients |  |
| MMWR 2020 (USA) [89] | All age | 4,226 | R | Age ≥ 65 vs <65= 8.84, 4.23- 18.48 |  | Age ≥ 65 vs <65= 2.62, 1.81-3.78 |

CS= Cross-sectional

ICU=intensive care unit

R=Retrospective

P: Prospective

HR= hazards ratio
