## Supplemental Table 4 for "Risk factors for severe illness and death in COVID-19: a systematic review and meta-analysis"

| S4 Table. Risk of bias and quality of the studies included in the review | | | | | | | |
| --- | --- | --- | --- | --- | --- | --- | --- |
| **References** | **Selection bias** | **Design** | **Confounders** | **Blinding (detection and reporting bias)** | **Data collection** | **Withdrawals and dropouts** | **Overall quality** |
| Aggarwal et al. 2020 [26] | 2 | 2 | 1 | 2 | 1 | 3 | 2 |
| Al-Rousan, 2020 [27] | 1 | 3 | 3 | 2 | 3 | 2 | 3 |
| Bezzio et al. 2020 [28] | 1 | 2 | 3 | 2 | 2 | 3 | 3 |
| Bi et al. 2020 [29] | 1 | 2 | 1 | 1 | 1 | 1 | 1 |
| Bi et al. 2020 [30] | 2 | 2 | 3 | 2 | 1 | 3 | 3 |
| MMWR, 2020 [31] | 1 | 3 | 3 | 2 | 1 | 2 | 3 |
| MMWR,2020 [24] | 1 | 3 | 3 | 2 | 1 | 2 | 3 |
| Chen et al. 2020 [32] | 2 | 2 | 1 | 2 | 2 | 1 | 1 |
| Colaneri et al. 2020 [33] | 2 | 2 | 3 | 3 | 1 | 1 | 3 |
| Deng G et al., 2020 [5] | 1 | 3 | 3 | 3 | 3 | 2 | 3 |
| Deng et al, 2020 [34] | 2 | 2 | 1 | 3 | 1 | 1 | 2 |
| Du et al. 2020b [35] | 2 | 2 | 1 | 2 | 1 | 1 | 1 |
| Du et al., 2020a [10] | 2 | 2 | 3 | 2 | 1 | 3 | 3 |
| Dudley et al. 2020 [36] | 1 | 3 | 3 | 3 | 3 | 3 | 3 |
| Feng et al. 2020 [37] | 2 | 2 | 1 | 2 | 2 | 1 | 1 |
| Gong et al., 2020 [38] | 2 | 2 | 3 | 3 | 2 | 3 | 3 |
| Grasselli et al.2020 [39] | 2 | 2 | 1 | 2 | 2 | 3 | 2 |
| Guan et al., 2020a [6] | 1 | 2 | 1 | 2 | 2 | 3 | 2 |
| Guan et al, 2020b [8] | 2 | 2 | 3 | 2 | 2 | 3 | 3 |
| Guo et al., 2020 [40] | 1 | 2 | 1 | 1 | 2 | 3 | 2 |
| Han et al., 2020 [41] | 1 | 2 | 1 | 3 | 1 | 3 | 3 |
| Hu et al., 2020 [42] | 1 | 2 | 1 | 3 | 1 | 1 | 3 |
| Ji et al., 2020 [43] | 1 | 2 | 3 | 2 | 1 | 1 | 2 |
| Kalligeros et al., 2020 [44] | 2 | 2 | 1 | 1 | 1 | 3 | 2 |
| Lei et al., 2020 [45] | 1 | 2 | 1 | 1 | 1 | 1 | 1 |
| Leung et al., 20202 [46] | 2 | 2 | 1 | 1 | 3 | 2 | 2 |
| Li et al., 2020 [47] | 2 | 2 | 1 | 2 | 1 | 3 | 2 |
| Li et al., 2020 [7] | 2 | 2 | 1 | 3 | 1 | 3 | 3 |
| Lian et al., 2020 [48] | 1 | 2 | 3 | 1 | 1 | 3 | 3 |
| Liu et al., 2020 [50] | 2 | 2 | 1 | 1 | 1 | 1 | 1 |
| Mehra et al., 2020 [51] | 2 | 2 | 1 | 1 | 1 | 1 | 1 |
| Mehta et al., 2020 [52] | 2 | 2 | 1 | 2 | 1 | 3 | 2 |
| Middeldrop et al., 2020 [53] | 2 | 2 | 1 | 2 | 2 | 1 | 1 |
| Nikpouraghdam et al., 2020 [54] | 2 | 2 | 1 | 1 | 1 | 3 | 2 |
| Pei et al., 2020 [55] | 2 | 2 | 1 | 2 | 1 | 1 | 1 |
| Pereira et al., 2020 [56] | 2 | 2 | 1 | 3 | 1 | 1 | 2 |
| MMWR, 2020 [90] | 1 | 3 | 3 | 2 | 1 | 3 | 3 |
| Qin et al., 2020 [57] | 2 | 2 | 1 | 2 | 1 | 3 | 2 |
| Qu et al., 2020 [58] | 2 | 3 | 1 | 3 | 1 | 2 | 3 |
| Richardson et al., 2020 [59] | 2 | 3 | 3 | 2 | 1 | 2 | 3 |
| Simonnet et al., 2020 [61] | 2 | 2 | 1 | 3 | 1 | 3 | 3 |
| Sun et al., 2020 [62] | 2 | 3 | 3 | 2 | 1 | 2 | 3 |
| Sun et al., 2020 [63] | 2 | 2 | 1 | 3 | 1 | 2 | 2 |
| Tang et al., 2020 [64] | 2 | 2 | 1 | 3 | 1 | 1 | 2 |
| Tedeschi et al., 2020 [65] | 2 | 2 | 1 | 3 | 3 | 3 | 3 |
| Tian et al., 2020 [66] | 2 | 2 | 3 | 2 | 1 | 1 | 2 |
| To et al., 2020 [67] | 2 | 2 | 3 | 2 | 1 | 3 | 3 |
| Verity et al., 2020 [68] | 2 | 3 | 1 | 3 | 3 | 2 | 3 |
| Wan et al., 2020 [69] | 2 | 2 | 3 | 3 | 1 | 1 | 3 |
| Wang et al., 2020 [70] | 2 | 2 | 1 | 3 | 1 | 1 | 2 |
| Wang et al., 2020 [71] | 2 | 3 | 1 | 3 | 1 | 2 | 3 |
| Wang et al., 2020 [72] | 2 | 2 | 1 | 2 | 1 | 1 | 1 |
| Wei et al, 2020 [73] | 2 | 2 | 1 | 1 | 1 | 3 | 2 |
| Wu et al.,2020 [74] | 2 | 2 | 1 | 3 | 1 | 1 | 2 |
| Yan et al., 2020 [75] | 2 | 2 | 1 | 3 | 1 | 1 | 2 |
| Yang G et al., 2020 [76] | 2 | 2 | 1 | 2 | 1 | 1 | 1 |
| Yang R et al., 2020 [77] | 2 | 2 | 3 | 3 | 1 | 1 | 3 |
| Yang X et al., 2020 [78] | 2 | 3 | 1 | 2 | 1 | 2 | 2 |
| Yao et al., 2020 [79] | 2 | 2 | 1 | 2 | 1 | 1 | 1 |
| Zhang R et al., 2020 [83] | 2 | 2 | 3 | 2 | 1 | 3 | 3 |
| Zou et al.2020 [86] | 2 | 2 | 3 | 3 | 1 | 1 | 3 |
| Zhang et al. 2020 [82] | 2 | 2 | 3 | 3 | 1 | 3 | 3 |
| Zhang et al. 2020 [84] | 2 | 2 | 1 | 3 | 1 | 3 | 3 |
| Zhang et al.2020 [85] | 2 | 1 | 1 | 3 | 1 | 2 | 2 |
| Zhang et al. 2020 [81] | 2 | 2 | 3 | 3 | 1 | 3 | 3 |
| Yu et al. 2020 [80] | 1 | 3 | 1 | 2 | 1 | 2 | 2 |
| Hu et al., 2020 [9] | 2 | 2 | 1 | 1 | 1 | 1 | 1 |
| Shi et al., 2020 [60] | 2 | 3 | 1 | 3 | 3 | 3 | 3 |
| Chen R et al., 2020 [87] | 1 | 2 | 1 | 1 | 1 | 1 | 1 |
| Dong Y et al., 2020 [88] | 2 | 2 | 3 | 3 | 3 | 3 | 3 |
| MMWR 2020 [89] | 1 | 3 | 3 | 2 | 1 | 2 | 3 |
